# Improving local authority financial support services for users with complex health needs: a mixed-method economic evaluation of Social Navigators in South Tyneside, UK

**DOI:** 10.1101/2024.11.07.24316923

**Authors:** Peter van der Graaf, Andrew McCarthy, Murali Subramanian, Bronia Arnott, Sarah Lee, Dilupa Samarakoon, Jo Gray, Angela Bate

## Abstract

**Objectives:** Applying social prescribing to non-clinical areas such as mental health, and underlying causes including financial hardship, is essential to support integrated care across the UK health and care system. There is inconclusive evidence on effectiveness of these services with a need for more mixed-methods research to understand if and how link worker roles improve outcomes for patients in communities. Our study aimed to evaluate the impact of a Social Navigators (SN) service in South Tyneside on the health and wellbeing of users who experience financial hardship and present with complex health needs.

**Design:** mixed methods study combing secondary analysis of service data with semi-structured interviews, conducted by peer researchers, and a Social Return On Investment analysis that matched service data with health economic indicators from the UK Social Value Bank.

**Setting:** metropolitan borough of South Tyneside, UK (population n=151,133), ranked 3^rd^ for employment, 13^th^ for income, and 15^th^ for health in term of most deprived areas out of 326 UK authorities.

**Participants:** 330 service users who engaged with the service between 2021-23. Most users are vulnerable with two-thirds being economically inactive, the majority earning less than £10,000, and 86% suffering with one or more health issues, with mental ill health being most common (58%). Interviews with15 service users opportunistically sampled from service database.

**Intervention:** Social Navigators working with residents who experience frequent financial hardship to identify and address underlying causes, by increasing their access to advice, health, employment and financial services, and building their skills and confidence in order to reduce health inequalities and dependency on crisis intervention.

**Results:** Our findings demonstrate clear value for money with a £3 social return for every £1 invested in the service with a positive return confirmed in sensitivity analysis. SN were able to improve service users’ confidence, with statistically significant changes across all eight confidence-related outcomes, and helped them to access other advice and financial services. This resulted in one-off financial gains (on average £1,237) and annual financial gains (on average £1,703) for service users. The interviews identified that relieving financial burden and stress improved the quality of life for and mental wellbeing of users as a result of their involvement with the service.

**Conclusions:** SN break the cycle of multiple visits to crisis teams by building trusting relationships and providing emotional and practical support, while being responsive to the service users’ needs. They play a key intermediary role in integrated care systems that is unique in its focus on the wider determinants of health and financial hardship, advocating for service users without time limits, and navigating the complexities of the system across local government. There is a need for better signposting and joining up of services to achieve a more whole systems approach to enhancing health & well-being in the community, while supporting the mental wellbeing of SN.

**Article Summary:** *Strengths and limitations of this study:*

- This is the first-mixed methods evaluation of social prescribing in the UK, focusing on the link between financial hardship and mental health.
- Applying a mixed-methods design allowed for combing local service monitoring data with national survey data to perform a Social Return on Investment analysis.
- Adding insights from service users through qualitative interviews granted the researchers insights in what outcomes mattered most to them and illuminated the mechanisms that they felt contributed to those outcomes.
- Using peer researchers to collect data from service users allowed for richer data collection through existing trusted relationships, while potential bias was checked through triangulation of different data sources.
- Closing gaps in local data collection, including longer-term follow-up data, and aligning data collection to national survey data would allow for more robust and less conservative SROI analysis.

## Introduction

Financial difficulties are a common cause of stress and anxiety and drastically reduce recovery rates for common mental health conditions. The impact on people’s mental and physical health can be particularly severe if they resort to cutting back on essentials, such as heating and eating, and there is a strong link between problem debt and suicide. Stigma around debt can also mean that people struggle to ask for help and may become isolated (1).

Tackling these complex and long-term health problems requires an extensive holistic approach not possible in routine primary care. Social prescription has been suggested as a potential intervention for addressing financial hardship by taking into account physical and mental health, and social and economic issues (2).

Social prescribing is a broad concept that has been applied across a range of contexts for a range of outcomes using a variety of models and activities (3, 4). This heterogeneity has made evaluation of interventions challenging with no agreed definition of social prescribing. However, it is generally understood to be a process that enables health care professionals to refer people to local, non-clinical services to support their health and wellbeing (5). This typically involves a link worker, also known as a community connector or navigator, who works with the individual to identify their needs, coproduce goals and connect them to resources in their community (6). Considerable funding has been committed by Central Government and the NHS to create these roles. For example, NHS Long Term Plan aimed to create 1,000 social prescribing link workers in primary care networks by 2020/2021 (7), while Government granted a further £5 million of funding for social prescribing to support COVID-19 recovery in recent years (8).

In spite of rapidly growing attention and funding for social prescription in the UK, its evidence base remains inconclusive. Existing evaluations have been criticised for lacking rigour with weaknesses in study designs and reporting of results due to a lack of comparative controls and standardised assessments, short duration, missing data and single point follow-up (9, 10).

There is limited evidence on the effectiveness of social prescribing roles, with most studies having been conducted in clinical settings, such as primary care ((11) substance use disorder treatment (12), support interventions for people living with HIV (13) and cancer communication (14)). In addition, these studies present inconclusive evidence on the impact of these roles on supporting patients with the challenges of treatment and maintain long-term recovery, although some navigators were effective in providing personalised guidance and support in areas such as medication adherence, disease disclosure, and accessing healthcare services.

The small number of studies in non-clinical areas involving community health and social workers only provide low quality evidence in the form of commentaries (15, 16) or untested toolkits (17), e.g. for training student volunteers to serve as community resource navigators. However, qualitative studies (18) suggest that link workers are able to support lifestyle changes by participants, and identify factors that facilitate progress towards intended goals, by giving patients the confidence, motivation, connections, knowledge and skills to manage their own well-being, thereby reducing their reliance on GPs (19).

Elliott et al. (20) conducted a realist review to understand how and why social prescribing evaluations work or do not work. Their recommendations outline an evaluation framework to improve methodological robustness of designs by applying mixed methods to capture the impact of social prescribing at multiple levels. This should involve an iterative design with local stakeholders actively involved in all stage of the research design, conduct and dissemination process, including members of the public, and triangulation of findings from multiple data sources and different perspectives to generate a more in-depth, nuanced understanding of social prescribing, how it works, for whom and in what context.

However, this framework remains untested with an urgent need to apply to non-clinical settings given the focus of social prescribing on addressing the wider determinants of health (21) and unique challenges of working across services in different systems, building relationships between different professions and organisations, and coordinating various intertwined working processes (4).

Therefore, this study presents the findings of a mixed-methods evaluation of a social prescribing service based within a local authority that works across different support services for people experiencing financial hardship while presenting with complex health and social needs. We will illustrate how this framework can be practically applied and demonstrate how this application enables new insights on the boundary spanning role of Social Navigators in integrated care systems.

### Introducing the intervention

The South Tyneside Social Navigator service was established in 2021 by South Tyneside Council (STC) South Tyneside Homes (STH) to addresses a gap in their support system where people in the Borough return multiple times for support from the crisis team without improving their financial circumstances. Service users also demonstrate complex additional health and social needs, which continuously impact on their ability to be financially stable.

Four Social Navigators (SN) were recruited to work inclusively and in a person-centred way with service users who presented most often to the Council’s welfare services. SN have an outreach role, meeting people on a 1:1 basis wherever they feel most comfortable in community settings. SN and service users work together for as long as needed, which can be up to six months or longer depending on circumstances. The SN work with service users to identify and address the underlying causes of the repeating cycle of wider issues which lead to frequent financial hardship and instability. This includes housing and tenancy issues, social isolation and loneliness, communication difficulties, fuel poverty, debt problems, access to washing/hygiene facilitates, and access to health and welfare services (mental and physical health).

The service aims to improve confidence and skills of service users to seek appropriate and timely assistance, increase access to advice, health, employment and financial services, to ultimately reduce dependency on crisis intervention. The Social Navigator also support the “knitting” together of services across the Borough and have well established links to at least 34 different organisations in South Tyneside, as well as internal links across the council. This includes specialist welfare benefit and debt advisors within the welfare support team, the Local Welfare Provision Scheme and various community organisations and charities. Not only do the SN refer into these services, but they also receive referrals from other services and organisations. The SN reach out to those who have actively disengaged from other services, such as households where children are affected by frequently re-occurring financial hardship.

### Co-producing the study

We used evaluability assessment methods to develop the evaluation design (22, 23). Evaluability Assessment (EA) is a rapid, systematic, and collaborative way of deciding whether and how a programme or policy can be evaluated, and at what potential cost. We conducted two EA workshops with SN stakeholders, to gauge their understanding of how the SN service was intended to work, how it might lead to health outcomes and how it may be evaluated (see Supplementary file 1). Workshop participants included staff from South Tyneside Council, South Tyneside Homes, local third sector organisations, and three public members who have engaged with the social navigators.

To support data collection and interpretation, an embedded researcher, who was a senior member of the Social Navigators team and was located in South Tyneside Homes, was funded for 2 days per week. Her support was instrumental in gaining access to service monitoring data, Social Navigators and service users, and crucial for disseminating the findings with the Council for recommissioning of the service.

In addition, four Social Navigators were recruited and trained as peer researchers to support data collection given the vulnerability of the client group. As SN, they have built up trusting relationships with clients, enabling them to elicit more meaningful responses. We anticipated that working with peer researchers would also improve response rates and build research capacity among Council staff by developing their interview skills. SN were provided with two online training sessions (2 hours each) by the research team over a two-week period in December 2023. The training covered research methodologies and using qualitative methods, including exercises for peer researchers to conduct interviews, and explored sensitive topics, such as ethical issues in research and handling challenging participants. Peer researchers were also given the interview topic guide and provided feedback on the questions and their acceptability to service users.

As a result of these workshops, training and input from our embedded researcher, our study aimed to explore and quantify the health and wellbeing impact of the Social Navigator project in South Tyneside on clients to inform recommissioning and future development of the service.

## Methods

We applied a mixed method design consisting of three interlinked data sources to reduce bias through triangulation of different data sources. The study was approved by Northumbria University Faculty of Health and Life Sciences research ethics committee (Van der Graaf 2023-4058-4031). We used the SQUIRE guidelines for reporting on our data (24).

### Secondary analysis of service monitoring data

We analysed existing monitoring data collected by the SN since the start of the service. Monitoring data consisted of participant characteristics (age, sex, employment status, health problems), referral data and financial gains achieved through support and referrals. It also included baseline and follow-up data on eight scored outcome measures that were developed by the service on a Likert scale from 1 (least confident) to 10 (most confident): ability to ask for help, access to advice, health and financial services, ability to budget income, digital social and employment skills).

We analysed these data through a prospective observational cohort study which included 330 participants from September 2021 to December 2023. Participant characteristics, scored outcome measures, and financial gains were reported using mean ± SD, median (IQR), or number (%). The impact of the SN on service users’ confidence as measured by the scored outcomes was explored through comparison of unadjusted differences at baseline and follow-up. In addition, the Wilcoxon signed rank test was used as a non-parametric statistical test to assess whether population mean ranks differ between baseline and follow-up for each outcome, p<0.05 considered significant. The Wilcoxon signed rank test was used as differences between pairs of data are non-normally distributed (25). All analyses were undertaken using SPSS version 29.

### Semi-structured interviews with service users

Potential gaps in understanding and missing data from the analysis of service monitoring data informed semi-structured interviews with a purposeful sample of 15 clients to explore in more detail the perceived health and wellbeing benefits of the Social Navigator project. Clients were sampled from the existing monitoring data to represent different referral pathways, varying lengths of support received from SN and a range of social-economic characteristics (age, gender, marital status, income).

The four trained peer researchers undertook a total of 15 interviews^1^ Most interviews took place in person, at the home of the service users, with peer researchers working in pairs to support each other. For example, one peer researcher would ask the questions, while the other peer researcher took notes and prompted service users, when needed for responses. Interviews lasted between 15 and 45 minutes, with an average of 30 minutes. In some interviews, carers were presented to support interviewees, and these carers contributed to the data collection by prompting the person they cared for in the interviews in response to questions asked by the interviewers. All interviews were digitally recorded, with consent, on voice recorders provided by the research team.

The recordings of the interviews were transcribed verbatim, inputted and coded in NVivo 12, by a member of the research team (MPS). A thematic analysis approach was utilised to analyse the interview transcripts. This method enabled the systematic identification and interpretation of recurring patterns and themes within the data. Following the guidelines outlined by Braun and Clarke (2006), the analysis process involved organising and interpreting qualitative data to disclose meaningful themes.

Once all the peer researcher-led interviews were completed, the research team organised an in-person feedback session to learn about their experiences and sense check the findings from the interviews. This feedback session helped the research team to identify any discrepancies or gaps in the data and gain more insight in the role of SN and their experiences of working with service users.

### Health economic modelling of health and wellbeing outcomes

Based on findings from WP1 and 2, we completed a Social Return On Investment (SROI) analysis by matching data from the South Tyneside SN service monitoring database to the social wellbeing values from the HACT database and calculating a return ratio. This return ratio is based on calculating inputs, outputs, outcomes and social values for the SN service, with added sensitivity analyses to test robustness of the results.

We calculated the costs to run the service (inputs) as well as identifying and measuring all feasible impacts of the service (outputs). Costs to run the Social Navigator service for one year were provided by South Tyneside Council in ≥2024 prices. This included staffing costs (covering SN roles and management), equipment costs (including smartphone, laptop), travel costs for staff, and a contributory share of the cost of access to an annual subscription case report service.

The monetary value of the financial gains of service users were regarded as benefits. In order to obtain a social value, the scored outcome measures collected by the SN service were mapped against the outcomes from the HACT Social Value Calculator (26). This calculator uses well-being valuation on national surveys to isolate the effect of a factor on an individual’s well-being and to identify an equivalent amount of money required to increase wellbeing by the same amount would be (27). To obtain a social value in the base-case for a scored outcome, measured from 1 (low) to 10 (high), a service user had to score 5 or less at baseline and a 6 or higher at follow-up.

To limit overestimation of social return on investment, a deadweight loss was assigned to outcomes. A deadweight loss is defined as a measure of the amount of outcome that would have happened even if the SN service did not take place (28). For the base-case analysis the social values that were mapped to the HACT Social Value Calculator were assigned the deadweight values that were estimated in HACT Social Value Calculator. The financial gains were not assigned a deadweight loss value for the base case analysis, as it was assumed that the service users would not have received those financial gains without the SN.

Sensitivity analysis was conducted to explore the uncertainty and robustness of the SROI results. Different underlying assumptions of the base case evaluation were changed to explore the impact on the SROI result, using more conservative and restrictive assumptions to see whether a threshold value (e.g. where SROI ratio crosses from positive to negative) could be identified. For example, by increasing the level of deadweight loss (to 50% and 75% respectively) or by increasing the threshold needed (from 6 to 8) to obtain the social value for the scored outcomes. A separate sensitivity analysis was conducted in which the financial gains were excluded entirely with benefit only being derived from the social values from the scored outcome measures.

### Patient and Public Involvement statement

Public involvement is an integral part of our ways of working in PHIRST Fusion, both at programme and project level, tailored to the needs of the local authority and their stakeholders, including public members. For example, public members have taken part in our evaluability assessment workshops to co-produce the study design, they supported the delivery of projects by becoming peer researchers, they helped to mobilise findings through existing patient and public involvement networks, and they facilitated the co-design of targeted outputs (e.g. community postcards). At programme level, we have two public members in our Independent Advisory Group (IAG) who actively involved beyond annual meetings and provide input for project reports, contribute to papers, provide guidance for local engagement activities with communities, and review our project knowledge mobilisation plans. For this paper, we mapped our three interlinked work packages onto the evaluation framework for social prescribing initiatives developed by Elliott et al. (2022) to illustrate how we applied their recommendations in our study design (Supplementary file 2). The table demonstrates a good fit with all 15 recommendations, demonstrating robustness, accountability and resulting in an in-depth, nuanced understanding of the Social Navigators service, how it works, for whom and in what context. However, we diverted from recommendation 9 (remove the burden on link workers and use independent researchers to collect data at the appropriate time point). Social Navigators in South Tyneside were keen to be involved in the data collection and interpretation; their involvement greatly enhanced the access to and the quality of the data. We supported them in their role as peer researchers by providing training, equipment, recruitment and information materials and organising debrief meetings after interviews. The overall evaluation was conducted by independent researchers from PHIRST Fusion, removing the burden from the local authority. However, we extended recommendations 3 and 4 to include data collection with stakeholders to create more opportunities for co-production and translation into practice.

## Findings

We first present the findings from the social monitoring data analysis to describe the characteristics of the participants involved in the evaluation and financial gains they were able to access through their engagement with the service. We then outline the social benefits to service users by first describing the change in confidence-related outcomes measures from the service monitoring data, combined with findings from the semi-structured interviews with service users. Finally, we will put a financial figure on these social benefits by reporting on the results from our social return on investment analysis.

### Participants characteristics

Between October 2021 and December 2023 there were 330 participants who engaged with the SN. Initial enquires, those who contacted the service but did not utilise it, consisted of 103 service users (94 closed enquiries and 9 ongoing) were excluded from the analysis. For example, some service users who disengaged after initial support or only required limited support with no scored outcomes data collected. Of the 227 remaining participants with sufficient engagement, 26 were excluded due to having an ongoing open case with limited follow-up data to record meaningful changes. The final cohort consisted of 201 closed cases (Figure 1).

**Figure 1.**
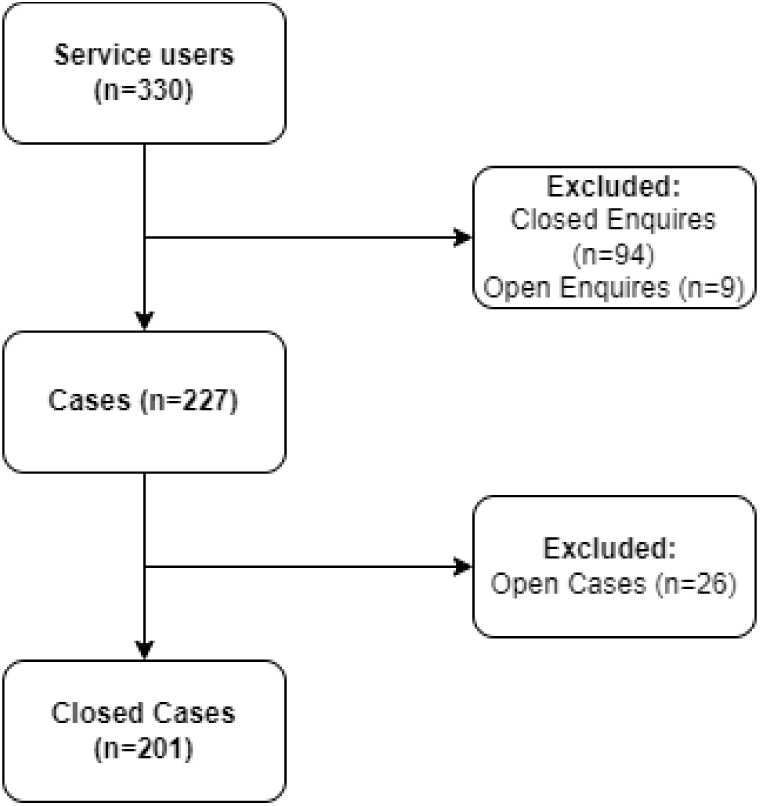
Sampling flow diagram

Most participants were female (n=132 (66%)) and aged between 30 and 60 (n=143 (71%)). Although many service users lacked data on ethnicity, most of those reporting were White (n=102 (51%)). Two thirds of service users were economically inactive, either being unfit for work (n=89 (44%)) or unemployed (n=45 (22%)), with most users earning less than £10,000 and very few earning more than £20,000 (n=12 (6%)). Health issues were reported by 173 (86%) of the service users with mental ill health being most commonly reported (n=117 (58%)). On average service users reported 1.41 health problems (± 0.754), with a maximum of 5. Other health problems (disabilities, long term health problems) ranged between 5%-15% for the cohort (Table 1).

**Table 1.**
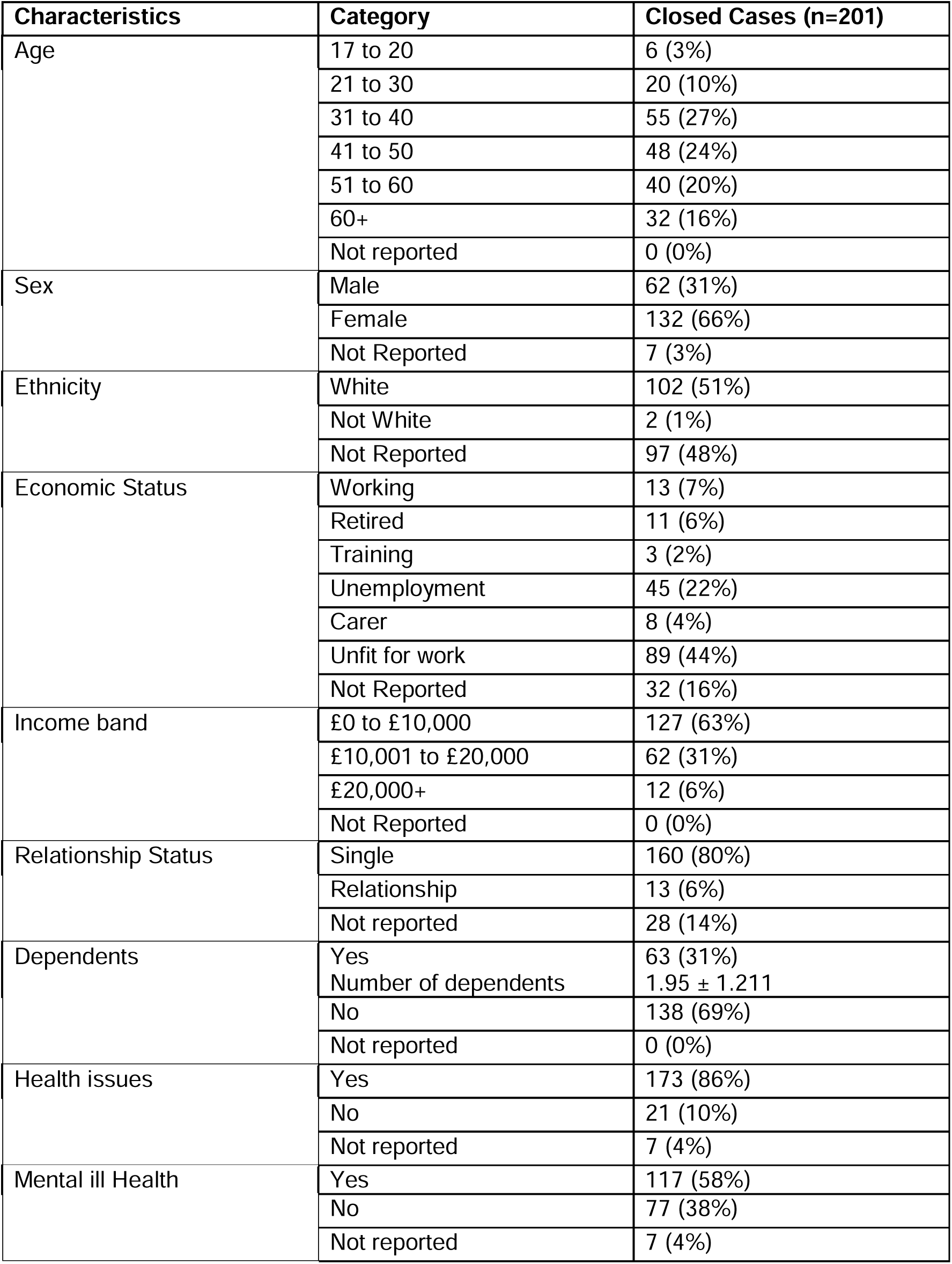
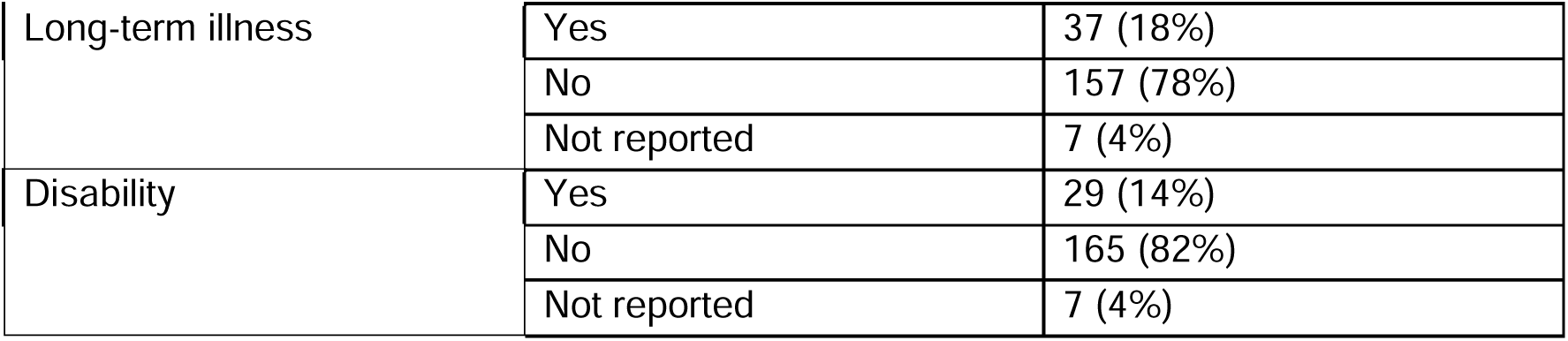
Participants baseline characteristics.

| Characteristics | Category | Closed Cases (n=201) |
| --- | --- | --- |
| Age | 17 to 20 | 6 (3%) |
|  | 21 to 30 | 20 (10%) |
|  | 31 to 40 | 55 (27%) |
|  | 41 to 50 | 48 (24%) |
|  | 51 to 60 | 40 (20%) |
|  | 60+ | 32 (16%) |
|  | Not reported | 0 (0%) |
| Sex | Male | 62 (31%) |
|  | Female | 132 (66%) |
|  | Not Reported | 7 (3%) |
| Ethnicity | White | 102 (51%) |
|  | Not White | 2 (1%) |
|  | Not Reported | 97 (48%) |
| Economic Status | Working | 13 (7%) |
|  | Retired | 11 (6%) |
|  | Training | 3 (2%) |
|  | Unemployment | 45 (22%) |
|  | Carer | 8 (4%) |
|  | Unfit for work | 89 (44%) |
|  | Not Reported | 32 (16%) |
| Income band | £0 to £10,000 | 127 (63%) |
|  | £10,001 to £20,000 | 62 (31%) |
|  | £20,000+ | 12 (6%) |
|  | Not Reported | 0 (0%) |
| Relationship Status | Single | 160 (80%) |
|  | Relationship | 13 (6%) |
|  | Not reported | 28 (14%) |
| Dependents | Yes | 63 (31%) |
|  | Number of dependents | 1.95 ± 1.211 |
|  | No | 138 (69%) |
|  | Not reported | 0 (0%) |
| Health issues | Yes | 173 (86%) |
|  | No | 21 (10%) |
|  | Not reported | 7 (4%) |
| Mental ill Health | Yes | 117 (58%) |
|  | No | 77 (38%) |
|  | Not reported | 7 (4%) |
| Long-term illness | Yes | 37 (18%) |
|  | No | 157 (78%) |
|  | Not reported | 7 (4%) |
| Disability | Yes | 29 (14%) |
|  | No | 165 (82%) |
|  | Not reported | 7 (4%) |

Reflecting on the interview with service users, SN highlighted the severe mental health issues faced by service users that they were trying to support:

> *“I mean when I first met her she was hysterical. She was going to give her kids up. Severe mental health problems, severe mental health problems.” [Social Navigator]*

> *“Well the work was stressing her out…and then something else had happened and she was just-she’s like ‘I can’t cope with this. It’s too much. It’s too much’.” [Social Navigator]*

### Financial gains

Financial gains were grouped into ongoing gains (annual gains assuming no changes in circumstances) and one-off gains. 133 (66%) of service users reported a financial gain after engaging with the SN service. Ongoing gains were achieved by 52 (25.9%) of the service users. The mean ± SD ongoing gain was £1703 ± £4,186. However, this is highly skewed with 7 participants receiving over £10,000, while 75% of participants received an ongoing gain of £634 or less.

Ongoing gains mostly consisted of gaining access to social security, with components of PIP forming most gains (n=56) comprising Personal Independence Payment (PIP) daily Living Enhanced rate (n=-14), PIP daily living standard rate (n=15), PIP mobility enhanced rate (n=14) PIP mobility standard rate (n=13). Council tax support (n=12) and University Credit (n=12) were also common gains. Table 2 shows a breakdown of ongoing annual gains sources and median gain per source.

**Table 2.** Financial Gains summary.

| Type of Gain | Measure | Closed Cases (n=201) |
| --- | --- | --- |
| Total one-off gains | Number of participants receiving gain | 133 (66%) |
| | Mean ± SD | £1,237 ± £844 |
|  | Median IQR | £180 (£0 to £992) |
| Total ongoing gains | Number of participants receiving gain | 52 (25.9%) |
| | Mean ± SD | £1703 ± £4,186 |
|  | Median IQR | £0 (£0 to £634) |

One-off gains were achieved by 137 (68%) participants, with a mean ± SD gain of £1,237 ± £844. Gains were highly skewed with a 50% of participants receiving a one-off gain of £180 or less, and 75% of participants receiving a one-off gain of £992 or less.

Common types of one-off gain included debt related gains (n=31), consisting of debt write off (n=15), debt managed (n=12) and debt relief order fee (n=4). Energy related gains (n=77) consisted of energy saving vouchers (n=34), energy savings (n=21) and energy saving lightbulbs (n=22). Household Support Fund (n=42) and Lucie’s Pantry Shopping (n=30) were also notable one-off gains. In addition, there were 44 instances of backdated payments and refunds related to PIP, universal credit, housing benefit. Gains consisted of a combination of government support and third sector support. Largest mean one-off gains came from Debt write offs and Backdated benefits (Table 2; see also Supplementary File 3).

### Confidence-related outcomes

173 (86.1%) service users completed the scored outcome questionnaire at baseline with 113 (55.4%) also completing a follow-up. Unadjusted mean scores at baseline were low across all eight questions, with the lowest response being for Seeking Employment (mean ± SD 1.05 ± 1.824) to the highest being Ability to ask for help (2.68 ± 1.513). There was an unadjusted mean ± SD gain of 3.212 ± 1.953 in service users’ confidence in ability to ask for help, following engagement with the SN. Confidence in accessing advice services (3.770 ± 2.32) and confidence in accessing financial services (2.142 ± 2.319) also saw large improvements. Limited gains were identified in confidence accessing health services (1.761 ± 2.028), confidence in being able to budget your income (1.814 ± 2.236), and social skills/socialisation (1.761 ± 1.938). With a small change in confidence in digital skills (0.690 ± 1.546), and confidence in seeking employment (0.513 ± 1.844) (Table 3).

**Table 3.** Scored Outcome measures.

| Scored Outcome | Baseline<br>(n=173) | Follow-up<br>(n=113) | Unadjusted<br>Difference (n=113) |
| --- | --- | --- | --- |
| Confidence in ability to ask for help | 2.68 ± 1.513 | 5.86 ± 1.968 | 3.212 ± 1.953 |
|  | 3 (2 to 3) | 6 (5 to 7) | 3 (2 to 4) |
| Confidence in accessing advice services | 2.35 ± 1.445 | 6.11 ± 2.102 | 3.770 ± 2.323 |
|  | 2 (1 to 3) | 6 (5 to 8) | 4 (3 to 5) |
| Confidence in accessing financial services | 1.38 ± 1.579 | 3.61 ± 2.986 | 2.142 ± 2.319 |
|  | 1 (0 to 2) | 4 (0 to 6) | 2 (0 to 4) |
| Confidence in accessing health services | 2.24 ± 2.228 | 4.12 ± 2.796 | 1.761 ± 2.028 |
|  | 1 (0 to 4) | 5 (1 to 6) | 1 (0 to 3) |
| Confidence in being able to budget your income | 2.19 ± 1.831 | 4.19 ± 2.610 | 1.814 ± 2.236 |
|  | 2 (1 to 3) | 4 (2 to 6) | 2 (0 to 3) |
| Confidence in digital skills | 1.81 ± 2.150 | 2.57 ± 2.474 | 0.690 ± 1.546 |
|  | 1 (0 to 3) | 2 (1 to 4) | 0 (0 to 1) |
| Confidence in seeking Employment | 1.05 ± 1.824 | 1.40 ± 2.359 | 0.513 ± 1.844 |
|  | 1 (0 to 1) | 1 (0 to 1) | 0 (0 to 0) |
| Confidence in social Skills/Socialising | 1.79 ± 1.903 | 3.54 ± 2.364 | 1.761 ± 1.938 |
|  | 1 (0 to 3) | 4 (1 to 5) | 1 (0 to 3) |
| *Data reported as mean ± SD or median (IQR).* |  |  |  |

However, there were statistically significant changes across all eight outcomes. The changes in scored outcomes demonstrate that the service can make significant improvements for this population. Not only in financial gains but also in improving their confidence and helping them to access other services.

### Peer interviews with service users and de-briefing with peer researchers

The changes in social benefits were confirmed by participants in the semi-structured interviews conducted by peer researchers. Service users described the impact of financial relief they achieved through assistance with debt management and accessing entitlements:

> *“The financial burden being taken off me is the biggest help for me as well as my partner” [P1]*

Relieving their financial burden improved quality of life for service users by becoming more able to leave their house and heat their homes:

> *“Well, I had a lot of debt on my gas meter, and you were able to get that on, because I didn’t have a supply for a year and a half.” [P2]*

In particular, users reported improvements in their mental well-being as a result of their involvement with the service. The emotional support and empowerment provided by the SN enhanced their mental health.

> *“He’s nowhere near as anxious as he was before…” [carer of P1]*

Participants credited SN focus on trust and relationship building as a key mechanism for achieving these benefits. By providing practical and emotional support, SN establish a sense of trust to build relationships with service users. This approach of offering help before asking for anything created a sense of reliability and goodwill, leading to a stronger and more trusting connection. Participants also valued the opportunity to confide in someone who understood their struggles.

> *“I felt like I could trust you a little bit more because you were giving before you were taking.” [P4]*

> *“And you listen, which is the main thing.” [P12]*

Building on this trust and relationship, service users felt confident to reach out to SN themselves whenever needed. Participants appreciated the accessibility and responsiveness of SN, feeling confident to reach out whenever needed.

> *“You always felt you could get in touch with her if you needed her for anything” [P5]*

Participants also highlighted the importance of tailored and personalised interactions to address their individual needs effectively.

> *“At first, the confusion of everything… trying to break that down and understand that, it was really daunting… But once that initially hit me and you kind of held my hand through it, I don’t think there was ever a reason for me not to fully engage.“*

The need for tailoring of support started at referral, with participants illustrating different referral pathways into the service, ranging from family members to various healthcare professionals, while other participants self-referred:

> *“I went to them to see if I could get them to help me get some discretionary housing benefit.” [P1]*

The need for non-judgemental support that is wide-ranging but consistent and reliable was echoed by the SN themselves when reflecting on the interview findings:

> *“Support is wide ranging. Even PIP assessments and being there when the doctor rings.” [Social Navigator]*

> *“It’s the consistency I find. A lot of them have been let down by other people in the past other services.” [Social Navigator]*

However, this tailored and personalised support for service users and the need to be always available and responsive came at a cost for the SN:

> *“There’s many a time I’ve come in-I mean I remember when I first started, I came in and said ‘I can’t do this job.’… because my poor old man who couldn’t get gas and electricity from anywhere all weekend. I was all ready to go round to his house and take money around.’” [Social Navigator]*

These experiences had a profound impact on the mental wellbeing of Social Navigators themselves which they recognised in reflecting on their work with service users. The emotional challenges faced by SN were exacerbated by the systematic challenges in trying to navigate between different services in South Tyneside and dealing with the complex pathways and systems.

Participants expressed frustration with systemic challenges and complexities, acknowledging that despite encountering negative experiences, they continue to navigate and work within the system to support their clients.

> *“The system works how the system works and sometimes it can treat you negatively. It’s not because of anything you’ve done it’s just the system.” [Social Navigator]*

> *“There’s a lot of things wrong with the system but you’ve got to-Work with it.” [Social Navigator]*

### Putting a value on social benefit: social return on investment analysis

We estimated a financial value for the social benefits, reported by service users and changes measured in confidence-related outcomes from the service monitoring data, by calculation a return ratio based on inputs, outputs, outcomes and social values for the SN service.

Staffing costs consisted of the largest portion of cost to running the SN service, with four SN having a total cost of £186,649 per annum (4 SN £38,000 per person, 1 manager £38,000). Additional costs included travel costs, which were estimated to be an additional £1,944 (assuming 90 miles per month at a cost of £0.45 per Social Navigator) and a SIM card cost of £480 (assuming £10 per month per Social Navigator).

Costs for equipment (e.g. laptop, smartphone etc.) were discounted over 5 years, with an estimated annualised economic cost of £350.73 per Social Navigator. Finally, there was an additional cost of £2,200, which was a proportional cost from the SN towards the annual subscription cost of a case report services used by South Tyneside for collecting data. The total annual cost of running the SN service was therefore £191,624. For the 23 months of service running (01 October 2021 to 30^th^ August 2023) there is an estimated total cost of £367,278.81.

By matching data from the South Tyneside SN service monitoring database to the social wellbeing values from the HACT database (Table 4), the total social value of benefits was estimated to be £1,090,987 in the base case scenario. Largest gains were obtained from confidence in accessing advice services (n=63, total gain £158,976), confidence in accessing health services (n=30, total gain £45,696), and confidence in social skills/socialising (n=16, total gain £295,378). Financial gains consisted of the additional gains to participants (Table 2).

**Table 4.** Social values of South Tyneside Social Navigators service.

| Stakeholder | Outcome | Financial Proxy | No. experiencing Outcome | Dead-weight | Attribution to Other Reasons | Drop-Off | Net Social Value |
| --- | --- | --- | --- | --- | --- | --- | --- |
| Service Users | Confidence in accessing advice services | £2,773 | 63 | 9% | 0% | 0% | £2,523 |
| Service Users | Confidence in social skills/ socialising | £3,209 | 30 | 11% | 0% | 0% | £2,856 |
| Service Users | Confidence in accessing health services | £12,623 | 16 | 22% | 0% | 0% | £9,846 |
| Service Users | Financial Gains | Monetary value of gains of participant | 136 | 0% | 0% | 0% | Monetary value of gains of participant |

The base case social return on investment was calculated by dividing total benefit by total cost, resulting in an estimated ratio of £2.97 of social value gained for every £1 spent. This suggests that there was a net benefit to South Tyneside council.

### Sensitivity analysis

Increasing the deadweight loss to 50% for the scored outcome measures, resulted in a lower estimated SROI ratio of 2.43:1, with an increase to (a very high conservative estimate of) 75% resulting in a further ratio reduction to 2.02:1. Both scenarios still resulted in positive social value.

Decreasing the amount of service users who gained social value from the scored outcomes (by using a higher inclusion threshold) resulted in a revised SROI ratio of 1.96:1. Moreover, excluding the financial gains to service users resulted in a much reduced but still positive SROI ratio of 1.36:1 (See Table 5). In all, the sensitivity analysis reduced the SROI ratios but in all cases that was a positive ratio with the SN providing a net SROI. This means that the SROI analysis are robust and that the return ratio is likely to be an underestimate.

**Table 5:**
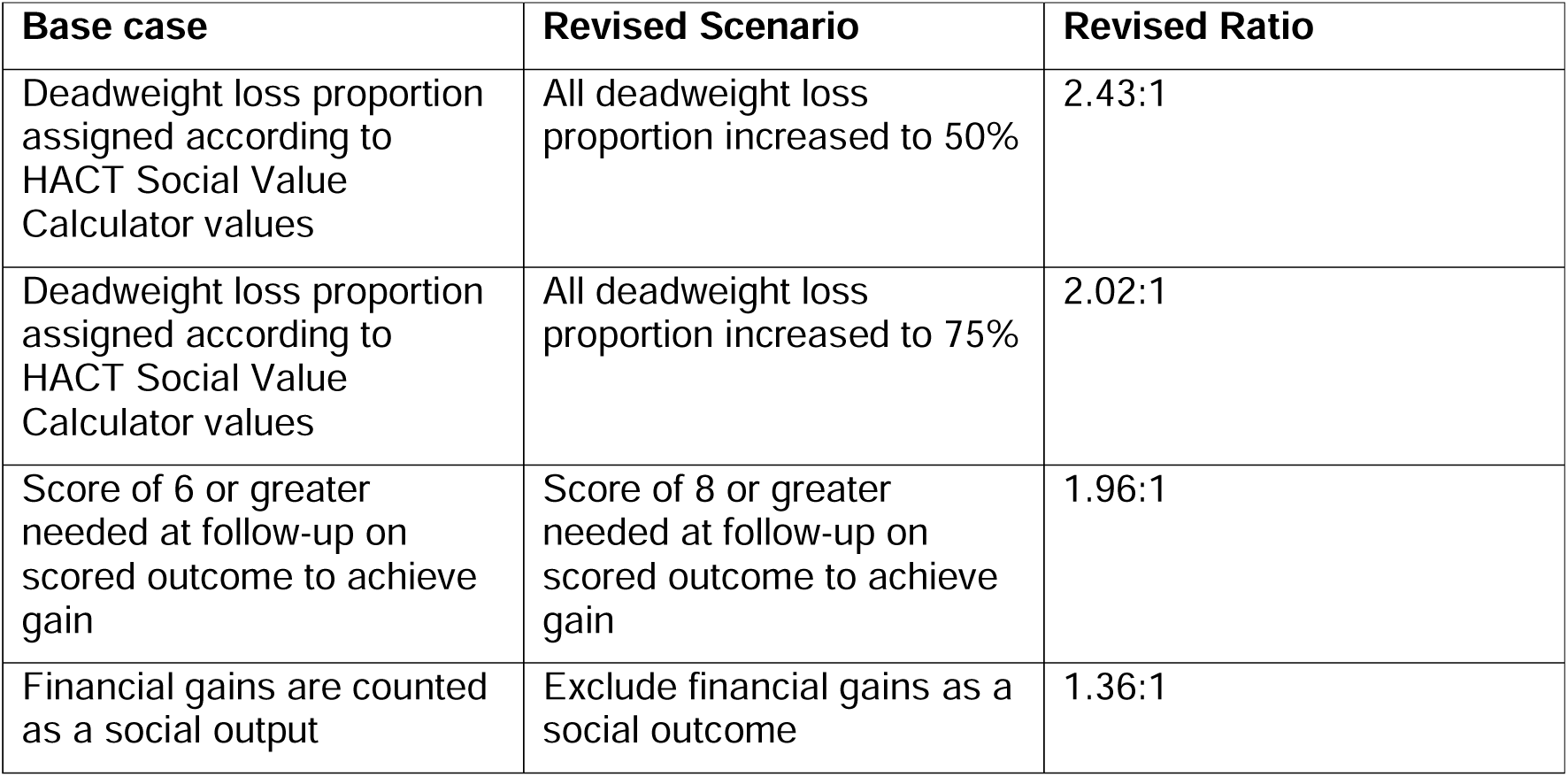
Sensitivity analysis; base case, revised scenarios and ratios.

| Base case | Revised Scenario | Revised Ratio |
| --- | --- | --- |
| Deadweight loss proportion assigned according to HACT Social Value Calculator values | All deadweight loss proportion increased to 50% | 2.43:1 |
| Deadweight loss proportion assigned according to HACT Social Value Calculator values | All deadweight loss proportion increased to 75% | 2.02:1 |
| Score of 6 or greater needed at follow-up on scored outcome to achieve gain | Score of 8 or greater needed at follow-up on scored outcome to achieve gain | 1.96:1 |
| Financial gains are counted as a social output | Exclude financial gains as a social outcome | 1.36:1 |

## Discussion

The changes in scored outcomes demonstrate that the social prescription services focused on non-clinical population targeting wider determinants of financial hardship can make significant improvements for this vulnerable population. The service monitoring data collected by the SN outline a vulnerable population with two thirds of service users being economically inactive, with most users earning less than £10,000, and 86% suffering with one or more health issues, with mental ill health being most commonly reported issue (58%).

This population has been falling between the cracks of service pathways in South Tyneside and several participants reflected on their previous negative experiences with other services. They returned multiple times for support from the crisis team, community funds and foodbanks, displaying complex additional health and social needs, which continuously impacted on their ability to be financially secure and stable. Their wide-ranging needs also significantly impacted on their ability to access services to get help for these needs, which in itself became a key factor in the cycle of re-occurring financial difficulties. They were faced by a difficult to navigate system, in which statutory support, e.g. from PIP or universal credit, can be time limited.

The data presented in this report suggests that SN can break this cycle and by building trusting relationships and providing emotional and practical support, while being responsive to the service users’ needs and available when they have needs. This support was followed up by tailored and personalised interactions, including help with accessing services and resources, such as filling out forms and making calls on behalf of the clients, who lacked sufficient digital and literacy skills. Their commitment and support were greatly valued by the service users and crucial in alleviating financial stress.

Service users benefited financially from the support of Social Navigators with two thirds (68%) receiving a one-off financial gain of on average £1,237 and over a quarter received an ongoing annual financial gain of on average £1,703. Service users also benefited socially, improving their confidence and access other services that further build their personal skills. There were statistically significant changes across all eight confidence-related outcomes.

While these gains might be perceived as small, they often made a big difference to the mental wellbeing and quality of life of service users. Relieving financial stress and burden helped them address other areas in their lives. The Social Return on Investment analysis put a financial figure on this support and estimated that the SN service had a positive social return of £3 for every £1 spent, suggesting that there was a clear net benefit to South Tyneside council.

Our findings suggests that the 1:1 support provided by SN elevated service users’ confidence in navigating other aspects of their lives, in line with other literature on social prescribing. For example, Bryant et al (2020) found that care navigators/link workers represented a vehicle for accruing social capital (e.g. trust, sense of belonging, practical support). They argue that this gives patients the confidence, motivation, connections, knowledge and skills to manage their own well-being, reducing their reliance on GPs.

The interviews with service users and SN debriefs highlighted the key intermediary role being played by the SN. They described their role as unique and distinct from social workers, as they bridge gaps in support systems by being human intermediaries and advocates for service users and navigating the frustrating complexities of the support system across the Borough. The focus of the social prescription service on non-clinical settings (local authority financial support service users) with a focus on the wider determinants of health as underlying causes for financial hardship, meant that SN had to elevate their role to become *system navigators*, working across multiple service within and outside the local authority, including local charities, businesses and community organisations.

This requires an additional skill set for the Social Navigators which is comparable to the skills of other boundary spanner roles, such as knowledge exchange brokers and embedded researchers. For example, Phipps and Morton (29) identified seven key skills for knowledge brokers: 1) being nimble, fleet footed (Mercury, the roman god with winged sandals), 2) enthusiastic (cheerleader), 3) creative (artist), 4) communicator, listener and supporter (therapist), 5) have courage (tightrope walker), 6) tact and negotiation skills (scales of justice),and 7) being tireless committed (athlete), resulting in an idealised picture that is almost impossible to achieve.

Similarly, Cheetham et al. (30) distinguished between five different roles that boundary spanners such as embedded researchers (ER) had to perform, highlighting the complexity, multi-facetedness and interconnectivity of these roles and the active work needed from individuals and organisations to put these roles into planning and maintaining them. ER acted as a sounding board, knowledge broker, facilitator, capacity builder and catalyst for change and improvement. For example, they needed develop and maintain effective working relationships with a diverse range of internal and external stakeholders to facilitate collaboration, co-production and networking, while working with people with complex mental health needs and long-term health conditions (which highlighted the value of service users’ stories and feedback in shaping services). Moreover, they needed to work with local partners to ensure that service/system transformation is catalysed using timely informal feedback (catalyst). This latter role seems particularly relevant for the Social Navigators roles but is perhaps also the most challenging, given the complexity of the local financial support system and lack of a map of available services to connect staff across services and share feedback between them.

While the authors of these studies argue that different qualities will be needed in different settings, nevertheless they agree that a basic ability across all qualities is important for successful boundary spanning roles and might require a team that can cover the qualities between them. They recommend a need for more training in these specialist qualities and explicit recruitment on these qualities when advertising for these boundary spanning roles.

This included a recognition of the challenges faced in system navigation roles. Feedback received from the Social Navigators after completing their peer researcher roles indicated that the diverse and demanding role put strain on the SN’s own health and wellbeing. When experiencing the hardship and emotional challenges faced by users and dealing with the difficulties in navigating between different services in the local authority, SN struggled to switch off from work and look after their own health and wellbeing. More support from Councils/ host organisations to deal with emotional challenges and maintain a healthy work-life balance is urgently needed. Access to organisational health and wellbeing resources, mentoring and coaching, and a longer-term contract to provide them with their own financial stability were recommended.

### Limitations

Limitations to this analysis are centred on data collection. Firstly, there was a large amount of missing data particularly around demographic information collected at baseline, and with scored outcome measures at follow-up. The missing data resulted in us being unable to use more complex evaluation methods, such as regression modelling. Included participant numbers would be too low, with techniques such as multiple imputation being unsuitable to overcome this issue. Furthermore, outcome data collected in this evaluation was focused on financial gains and the scored outcome measures. The scored outcome measures were focussed on service users ‘confidence in accessing services’ rather than whether services users accessed additional services (which could be costed) and outcomes of using those service (which could potentially be assigned a social value). Missing data and changes to collected outcome measures are something the SN are aware of because of their involvement in this evaluation and are looking to improve both response rates and what data is collected, with particular focus on mapping outcomes to HACT social values where possible. The missing data also limits the generalizability of the work beyond the local authority context of South Tyneside, which we hope the address in follow-up research with more complete and longer term data. We have made recommendations for improving the collection and use of routine data within the Council, some of which have already been implemented.

## Conclusions

Our evaluation of the Social Navigator (SN) service in South Tyneside strengthens the evidence base on social prescribing using a mixed methods design to demonstrate a net positive Social Return On Investment, which supported the recommissioning of the service and demonstrates the potential for spread to other UK local authorities. SN support those in the lowest socio-economic groups, who have little confidence in navigating services and who are at risk of falling through the cracks of current statutory service provision. SN break the cycle of multiple visits to crisis teams by building trusting relationships and providing emotional and practical support, while being responsive to the service users’ needs and available when they have needs. They play a key intermediary role in integrated care systems that is unique in its focus on the wider determinants of health and financial hardship, advocating for service users without time limits, and navigating the complexities of the system across local government. There is a need for more training and support for these diverse and challenging system navigating roles, given the range of activities they need to perform, and skills sets that are required to be successful. Repeating the SROI with more complete and better matched data over time could more powerfully demonstrate the difference SN are making, not only to service users but also to other services and the support system across local authorities.

## Supporting information

checklist

## Data Availability

All data produced in the present study are available upon reasonable request to the authors

## Declarations

### Ethics approval and consent to participate

Our study complies with the appropriate national research ethics process and R&D and governance processes. The study was externally reviewed and approved by Northumbria University Faculty of Health and Life Sciences research ethics committee (Van der Graaf 2023-4058-4031). Data are not publicly available, as it includes individual level data that allows for the identification of participants in our study, but are available from the corresponding author on reasonable request.

### Consent for publication

Individual written informed consent for participation in the research and for publication of anonymised data was obtained from each respondent in our study.

### Data statement

The datasets, which include anonymised interview transcripts and a description of our coding trees, are available from the corresponding author on reasonable request. They will be made available in the Northumbria University’s Research Repository. The final evaluation report with more details on the findings can be accessed from the NIHR PHIRST website: https://phirst.nihr.ac.uk/wp-content/uploads/2024/09/South-Tyneside-Social-Navigators-Evaluation-Final-Report.pdf.

### Author statement

South Tyneside Council conceived the idea for the study, PvdG, MS and BA developed the study design with input from all authors. Qualitative data collection and analysis was undertaken by MS. Quantitative data analysis was undertaken by AMcC, DS, JG and AB. Data interpretation was supported by all authors. The paper was drafted by PvdG and was commented on by all authors, who approved the final version.

## Acknowledgements

We would like to thank the Social Navigators who gave up their time to contribute to this study and without whose input and support in the data collection, analysis and interpretation, this study would not have been possible: Deborah Harper, Abigail Berry, James Auty and James Fellows.

## Footnotes

1 One of the peer researchers was not able to conduct any interview due to sick leave in the data collection period.

