## Supplementary material for "Improving local authority financial support services for users with complex health needs: a mixed-method economic evaluation of Social Navigators in South Tyneside, UK": checklist

### Reporting checklist for quality improvement in health care.

Based on the SQUIRE guidelines.

#### Instructions to authors

Complete this checklist by entering the page numbers from your manuscript where readers will find each of the items listed below.

Your article may not currently address all the items on the checklist. Please modify your text to include the missing information. If you are certain that an item does not apply, please write "n/a" and provide a short explanation.

Upload your completed checklist as an extra file when you submit to a journal.

In your methods section, say that you used the SQUIREreporting guidelines, and cite them as:

Ogrinc G, Davies L, Goodman D, Batalden P, Davidoff F, Stevens D. SQUIRE 2.0 (Standards for QUality Improvement Reporting Excellence): revised publication guidelines from a detailed consensus process

|  |  | Reporting Item | Page Number |
| --- | --- | --- | --- |
| **Title** |  |  |  |
|  | [#1](https://www.goodreports.org/reporting-checklists/squire/info/#1) | Indicate that the manuscript concerns an initiative to improve healthcare (broadly defined to include the quality, safety, effectiveness, patientcenteredness, timeliness, cost, efficiency, and equity of healthcare) | 1 |
| **Abstract** |  |  |  |
|  | [#02a](https://www.goodreports.org/reporting-checklists/squire/info/#02a) | Provide adequate information to aid in searching and indexing | 3 |
|  | [#02b](https://www.goodreports.org/reporting-checklists/squire/info/#02b) | Summarize all key information from various sections of the text using the abstract format of the intended publication or a structured summary such as: background, local problem, methods, interventions, results, conclusions | 2-3 |
| **Introduction** |  |  |  |
| Problem description | [#3](https://www.goodreports.org/reporting-checklists/squire/info/#3) | Nature and significance of the local problem | 4 |
| Available knowledge | [#4](https://www.goodreports.org/reporting-checklists/squire/info/#4) | Summary of what is currently known about the problem, including relevant previous studies | 4-5 |
| Rationale | [#5](https://www.goodreports.org/reporting-checklists/squire/info/#5) | Informal or formal frameworks, models, concepts, and / or theories used to explain the problem, any reasons or assumptions that were used to develop the intervention(s), and reasons why the intervention(s) was expected to work | 4-5 |
| Specific aims | [#6](https://www.goodreports.org/reporting-checklists/squire/info/#6) | Purpose of the project and of this report | 5 |
| **Methods** |  |  |  |
| Context | [#7](https://www.goodreports.org/reporting-checklists/squire/info/#7) | Contextual elements considered important at the outset of introducing the intervention(s) | 5-6 |
| Intervention(s) | [#08a](https://www.goodreports.org/reporting-checklists/squire/info/#08a) | Description of the intervention(s) in sufficient detail that others could reproduce it | 5-6 |
| Intervention(s) | [#08b](https://www.goodreports.org/reporting-checklists/squire/info/#08b) | Specifics of the team involved in the work | 6 |
| Study of the Intervention(s) | [#09a](https://www.goodreports.org/reporting-checklists/squire/info/#09a) | Approach chosen for assessing the impact of the intervention(s) | 7-10 |
| Study of the Intervention(s) | [#09b](https://www.goodreports.org/reporting-checklists/squire/info/#09b) | Approach used to establish whether the observed outcomes were due to the intervention(s) | 7-10 |
| Measures | [#10a](https://www.goodreports.org/reporting-checklists/squire/info/#10a) | Measures chosen for studying processes and outcomes of the intervention(s), including rationale for choosing them, their operational definitions, and their validity and reliability | 7-10 |
| Measures | [#10b](https://www.goodreports.org/reporting-checklists/squire/info/#10b) | Description of the approach to the ongoing assessment of contextual elements that contributed to the success, failure, efficiency, and cost | 7-10 |
| Measures | [#10c](https://www.goodreports.org/reporting-checklists/squire/info/#10c) | Methods employed for assessing completeness and accuracy of data | 8-10 |
| Analysis | [#11a](https://www.goodreports.org/reporting-checklists/squire/info/#11a) | Qualitative and quantitative methods used to draw inferences from the data | 7-10 |
| Analysis | [#11b](https://www.goodreports.org/reporting-checklists/squire/info/#11b) | Methods for understanding variation within the data, including the effects of time as a variable | 7-10 |
| Ethical considerations | [#12](https://www.goodreports.org/reporting-checklists/squire/info/#12) | Ethical aspects of implementing and studying the intervention(s) and how they were addressed, including, but not limited to, formal ethics review and potential conflict(s) of interest | 6-7/ 24 |
| **Results** |  |  |  |
|  | [#13a](https://www.goodreports.org/reporting-checklists/squire/info/#13a) | Initial steps of the intervention(s) and their evolution over time (e.g., time-line diagram, flow chart, or table), including modifications made to the intervention during the project | n/a. We conducted the evaluation to inform future development of the intervention. |
|  | [#13b](https://www.goodreports.org/reporting-checklists/squire/info/#13b) | Details of the process measures and outcome | 12-17 |
|  | [#13c](https://www.goodreports.org/reporting-checklists/squire/info/#13c) | Contextual elements that interacted with the intervention(s) | 15-16 |
|  | [#13d](https://www.goodreports.org/reporting-checklists/squire/info/#13d) | Observed associations between outcomes, interventions, and relevant contextual elements | 12-17 |
|  | [#13e](https://www.goodreports.org/reporting-checklists/squire/info/#13e) | Unintended consequences such as unexpected benefits, problems, failures, or costs associated with the intervention(s). | 15-16/ 20 |
|  | [#13f](https://www.goodreports.org/reporting-checklists/squire/info/#13f) | Details about missing data | 20 |
| **Discussion** |  |  |  |
| Summary | [#14a](https://www.goodreports.org/reporting-checklists/squire/info/#14a) | Key findings, including relevance to the rationale and specific aims | 17-19 |
| Summary | [#14b](https://www.goodreports.org/reporting-checklists/squire/info/#14b) | Particular strengths of the project | 20 |
| Interpretation | [#15a](https://www.goodreports.org/reporting-checklists/squire/info/#15a) | Nature of the association between the intervention(s) and the outcomes | 17-19 |
| Interpretation | [#15b](https://www.goodreports.org/reporting-checklists/squire/info/#15b) | Comparison of results with findings from other publications | 18-19 |
| Interpretation | [#15c](https://www.goodreports.org/reporting-checklists/squire/info/#15c) | Impact of the project on people and systems | 17-19 |
| Interpretation | [#15d](https://www.goodreports.org/reporting-checklists/squire/info/#15d) | Reasons for any differences between observed and anticipated outcomes, including the influence of context | 15-16 |
| Interpretation | [#15e](https://www.goodreports.org/reporting-checklists/squire/info/#15e) | Costs and strategic trade-offs, including opportunity costs | 16 |
| Limitations | [#16a](https://www.goodreports.org/reporting-checklists/squire/info/#16a) | Limits to the generalizability of the work | 20 |
| Limitations | [#16b](https://www.goodreports.org/reporting-checklists/squire/info/#16b) | Factors that might have limited internal validity such as confounding, bias, or imprecision in the design, methods, measurement, or analysis | 20 |
| Limitations | [#16c](https://www.goodreports.org/reporting-checklists/squire/info/#16c) | Efforts made to minimize and adjust for limitations | 7/ 17 |
| Conclusion | [#17a](https://www.goodreports.org/reporting-checklists/squire/info/#17a) | Usefulness of the work | 21 |
| Conclusion | [#17b](https://www.goodreports.org/reporting-checklists/squire/info/#17b) | Sustainability | 21 |
| Conclusion | [#17c](https://www.goodreports.org/reporting-checklists/squire/info/#17c) | Potential for spread to other contexts | 21 |
| Conclusion | [#17d](https://www.goodreports.org/reporting-checklists/squire/info/#17d) | Implications for practice and for further study in the field | 21 |
| Conclusion | [#17e](https://www.goodreports.org/reporting-checklists/squire/info/#17e) | Suggested next steps | 21 |
| **Other information** |  |  |  |
| Funding | [#18](https://www.goodreports.org/reporting-checklists/squire/info/#18) | Sources of funding that supported this work. Role, if any, of the funding organization in the design, implementation, interpretation, and reporting | 24-25 |

None The SQUIRE 2.0 checklist is distributed under the terms of the Creative Commons Attribution License CC BY-NC 4.0. This checklist can be completed online using <https://www.goodreports.org/>, a tool made by the [EQUATOR Network](https://www.equator-network.org) in collaboration with [Penelope.ai](https://www.penelope.ai)
