## Supplementary File 1 for "Improving local authority financial support services for users with complex health needs: a mixed-method economic evaluation of Social Navigators in South Tyneside, UK"

South Tyneside Social Navigator Project Evaluability Assessment Report

**PHIRST Fusion**

April 2023

### Summary

The South Tyneside Social Navigators (SN) project is an intervention offered by South Tyneside Homes in South Tyneside since September 2021, funded to September 2023. It is an impartial advice team that serve all residents of the borough of South Tyneside regardless of their tenure. Social Navigators provide intensive support to residents experiencing financial wellbeing issues. SN was allocated to PHIRST Fusion in November 2022. This report describes an Evaluability Assessment (EA) of SN conducted between January and March 2023. The EA workshops established that the overall aim of the SN evaluation is to understand how and why improvement in financial gains for project participants lead to better health and wellbeing. A few evaluation design options were considered for this evaluation and the recommended option combines secondary data analysis of existing monitoring data with semi-structured follow up interviews and focus groups. A health economic work package will also model health and wellbeing outcomes.

### 1 Introduction

#### Background to the PHIRST scheme & Social Navigators project

This evaluation initiative is funded by the National Institute for Health and Care Research (NIHR) and undertaken by the Public Health Intervention Responsive Studies Team (PHIRST) Fusion team. The PHIRST Fusion team’s approach to evaluation follows a 5-step process: brokerage, work allocation, research, reporting & knowledge mobilisation, and continuous improvement, which includes evaluability assessment methodology and embedded research with local government practitioners. The South Tyneside Social Navigator Project (SN) was allocated to PHIRST Fusion in November 2022. This report describes an Evaluability Assessment of SN conducted between January and March 2023.

SN is an intervention offered by South Tyneside Homes in South Tyneside since September 2021, funded to September 2023. It is an impartial advice team that serve all residents of the borough of South Tyneside regardless of their tenure. Social Navigators provide intensive support to residents experiencing financial wellbeing issues. South Tyneside Council required an initial phase of evaluation to provide evidence to support local authority decision-making about continuation of funding.

#### Programme context

Financial difficulties are a common cause of stress and anxiety and drastically reduces recovery rates for common mental health conditions. The impact of people's mental and physical health can be particularly severe if they resort to cutting back on essentials, such as heating and eating, and there is a strong link between problem debt and suicide. Stigma around debt can also mean that people struggle to ask for help and may become isolated (Money and Mental Health Policy Institute 2021). South Tyneside Homes Welfare Support Team are an impartial advice team that serve all residents of the borough of South Tyneside regardless of their tenure. They also administer the Local Welfare Provision service in the borough on behalf of South Tyneside Council.

The welfare support team noted people returning multiple times for support from the crisis team, community funds and foodbanks. Despite repeated access to these services, their financial circumstances were not improving, and they were still financially dependent and not reaching financial stability. Service users who repeatedly used the crisis and welfare support services reported that they had complex additional health and social needs which continuously impacted on their ability to be financially secure and stable. These wide-ranging needs were not just housing or tenancy issues but included social isolation, communication difficulties, fuel poverty, benefit, debt and employment services, access to washing/hygiene facilitates, access to health and welfare services (mental and physical health), lack of skills (including cooking, home hygiene and selfcare). These needs also significantly impacted on people’s ability to access services to get help and thus perpetuate a cycle of re-occurring financial difficulties. This was reflected in the 2019 Indices for Multiple Deprivation where access to housing and services in South Tyneside was ranked as 266th out of 317 (where higher rank relate to higher levels of deprivation) (JSNAA 2021).

In 2021 the role of Social Navigators was established to work inclusively and in a person-centred way with our service users who present most often to our welfare services. These social navigators have an outreach role, meeting people on a 1:1 basis wherever they feel most comfortable in community settings. Social navigators work with service users for as long as needed (‘stick to them like glue’), which can be up to six months or longer depending on circumstances. The social navigators help service users to identify and address the underlying causes of the repeating cycle of wider issues which lead to frequent financial hardship and instability. The service aims to improve confidence and skills to seek appropriate and timely assistance, increase access to advice, health, employment, and financial services, which will reduce health inequalities and reduce financial exclusion and dependency on crisis intervention.

### 2 Rapid Review

There is an extremely limited literature that is applicable to this area of research. Below is a summary.

Gautam, D., Sandhu, S., Kutzer, K., Blanchard, L., Xu, J., Munoz, V.S., Dennis, E., Drake, C., Crowder, C., Eisenson, H. and Bettger, J.P., 2022. Training student volunteers as community resource navigators to address patients' social needs: A curriculum toolkit. Frontiers in Public Health, 10.

In this article, Gautam et al. describe a curriculum toolkit for training student volunteers to serve as community resource navigators, who can help address patients' social needs. The authors argue that addressing social determinants of health is an important aspect of healthcare, and student volunteers can play a vital role in this effort. The toolkit provides guidance on how to train volunteers in key skills such as communication, problem-solving, and resource navigation, and also includes case examples to help students apply these skills in real-world scenarios.

Wells, K.J., Dwyer, A.J., Calhoun, E. and Valverde, P.A., 2021. Community health workers and non-clinical patient navigators: A critical COVID-19 pandemic workforce. Preventive Medicine, 146, p.106464.

This commentary explores the role of community health workers and non-clinical patient navigators in responding to the COVID-19 pandemic. The authors argue that these workers, who are often trusted members of the community they serve, can play a crucial role in addressing health disparities and improving health outcomes during the pandemic. The article also provides examples of successful programs that have utilized community health workers and patient navigators to address COVID-19-related challenges, such as vaccine hesitancy and access to care. The authors suggest that investing in these types of workers can have significant benefits for both public health and healthcare systems.

Davis, T. C., Williams, M. V., Marin, E., Parker, R. M., & Glass, J. 2002. Health literacy and cancer communication. CA: A Cancer Journal for Clinicians, 52(3), 134-149.

This study examines the role of health literacy and social navigation in cancer communication. The study found that individuals with low health literacy face challenges in accessing and understanding cancer information and that social navigators, including healthcare providers and family members, can help address these challenges by providing personalized guidance and support.

Darnell, J.S., 2013. Navigators and assisters: two case management roles for social workers in the Affordable Care Act. Health & social work, 38(2), pp.123-126.

This commentary discusses the important roles of social workers in implementing the Affordable Care Act (ACA) through the Navigator and Assister programs. The Navigator program aims to provide guidance and support to individuals and families enrolling in health insurance plans through the ACA, while the Assister program focuses on helping individuals access health care services. The article highlights the skills and competencies that social workers bring to these roles, including case management, advocacy, and cultural competence. The author argues that social workers can play a critical role in ensuring that vulnerable populations have access to health care services under the ACA, and that the Navigator and Assister programs offer opportunities for social workers to engage in this work.

Systematic / Scoping Reviews:

Krulic, T., Brown, G. and Bourne, A., 2022. A Scoping Review of Peer Navigation Programs for People Living with HIV: Form, Function and Effects. AIDS and Behavior, pp.1-21.

This scoping review examined the role of social navigation as a support intervention for people living with HIV. The study found that social navigators, including peer mentors and community health workers, were effective in providing guidance and support in areas such as medication adherence, HIV disclosure, and accessing healthcare services.

Gormley, M.A., Pericot-Valverde, I., Diaz, L., Coleman, A., Lancaster, J., Ortiz, E., Moschella, P., Heo, M. and Litwin, A.H., 2021. Effectiveness of peer recovery support services on stages of the opioid use disorder treatment cascade: A systematic review. Drug and Alcohol Dependence, 229, p.109123.

This systematic review examined the role of peer navigators in substance use disorder treatment. The study found that social navigators, including peers, recovery coaches, and sponsors, were inconclusive in helping individuals with substance use disorders navigate the challenges of treatment and maintain long-term recovery.

A list of potential health and wellbeing outcomes and their measures, based on previous research that other members of the PHRIST team were aware of is also provided here.

Table 1 Health and wellbeing outcomes for STC Social Navigators project from previous research

| ***Outcomes*** | ***indicator*** | ***Used in research*** | ***Comments*** |
| --- | --- | --- | --- |
| Mental wellbeing | Shortened Warwick Edinburgh Mental Wellbeing Scale | a mixed methods study of successful components in an integrated wellness service in North East England ([Cheetham et al. 2018](https://bmchealthservres.biomedcentral.com/articles/10.1186/s12913-018-3007-z)) | In reality it was not administered to every service user and follow-up measures were often not reported. |
| Self-efficacy, enhanced self-esteem, and improved confidence and motivation | Qualitative data from interviews | [Cheetham et al. 2018](https://bmchealthservres.biomedcentral.com/articles/10.1186/s12913-018-3007-z) | provided specific information on lifestyle changes made by participants, and explored factors reported to facilitate progress towards intended goals. |
| Sustained relationships with link worker and social interactions with other people | Qualitative data from interviews | [Cheetham et al. 2018](https://bmchealthservres.biomedcentral.com/articles/10.1186/s12913-018-3007-z) |  |
| Social isolation | Qualitative data from interviews | [Cheetham et al. 2018](https://bmchealthservres.biomedcentral.com/articles/10.1186/s12913-018-3007-z) |  |
| Social value of wellbeing | HACT Wellbeing values | <http://hact.org.uk/publications/> (suggested by Cheetham et al. 2018) | Regression analysis to estimate the relationships between subjective wellbeing and the various outcome variables included in the value bank. Followed by wellbeing valuation method. This approach relies on a comparison between the change in wellbeing from the outcome to be valued with the change in wellbeing from income. The value of the outcome is then calculated as the marginal rate of substitution (MRS) between income and the outcome itself, expressed in monetary terms. |
| Behaviour change outcomes (i.e. whether client goals in relation to diet, physical activity, smoking or other behaviours, had been achieved) | Outcomes were recorded in online reporting system to estimate health gains in quality-adjusted life years (QALYs). | Used in evaluation of lay health workers in Durham ([Visram et al,. 2020](https://www.sciencedirect.com/science/article/pii/S0277953619306562?casa_token=sRuQAfouUnsAAAAA:WEYaZVdCT5OrKIQJm2U3wez-Gw592c4jj1eg464Gwan7CecQjpjtansku0GvQUqOPOH3kcOL21aK)), applying  a ‘[ready reckoner](https://www.building-leadership-for-health.org.uk/evaluating-behaviour-change/health-trainers-health-economics-behavioural-economics-new-media/)’/ economic model (Lister, 2010; updated in 2016 using 2014/15 values) | Model assessed health trainer performance in relation to service objectives and compare this to costs, based on assumptions drawn from published evidence of the short- and long-term impacts of behaviour change. Other activities, such as asset mapping (identifying the existing strengths and resources within target communities) and signposting (referring clients to other services or activities), were valued by comparing the costs and outcomes with broadly similar primary care interventions. The estimates were then adjusted to take into account impact on health inequalities by applying a factor derived from the Health England Leading Prioritisation (HELP) review, to reflect the value of targeting [disadvantaged groups](https://www.sciencedirect.com/topics/social-sciences/disadvantaged-group) ([Health England, 2009](https://www.sciencedirect.com/science/article/pii/S0277953619306562?casa_token=sRuQAfouUnsAAAAA:WEYaZVdCT5OrKIQJm2U3wez-Gw592c4jj1eg464Gwan7CecQjpjtansku0GvQUqOPOH3kcOL21aK" \l "bib15)). |
| Social capital | Scale questions on trust, sense of belonging, practical support | [Tierney et al. 2020](https://bmcmedicine.biomedcentral.com/articles/10.1186/s12916-020-1510-7) summarised in NIHR [evidence brief](https://evidence.nihr.ac.uk/alert/social-prescribing-could-empower-patients-to-address-non-medical-problems-in-their-lives/) | Social capital gives patients the confidence, motivation, connections, knowledge and skills to manage their own well-being, thereby reducing their reliance on GPs |
| Patient activation | patient activation measure to gauge how motivated and able someone is to manage their health | [Hibbard et al., 2004](https://onlinelibrary.wiley.com/doi/pdf/10.1111/j.1475-6773.2004.00269.x), used in Tierney et al. 2020: | People identified as activated on this measure appeared more likely to adopt healthy behaviours (e.g. diet and exercise) and to have less hospital use |
| Patient outcomes:   - improvements in general health and wellness such as, reduced unmet needs, improved mental health, and reduced co-morbidities. - improved self-efficacy, self-management or empowerment. - increased patient satisfaction with services - Increased access to care or better follow up care - Patient encounters and communication with primary care (increased visits, improved communication, more reviews, check-ins and/or goal setting conducted and links made to other providers).   increased employment, reduced financial stresses, improved insurance coverage. | | | Outcomes identified in a [scoping literature review](https://bmchealthservres.biomedcentral.com/articles/10.1186/s12913-017-2046-1#Sec6) of papers from Canada, the United States, the United Kingdom, Australia, New Zealand, and/or Western Europe published between January 1990 and June 2013 if they discussed navigators or navigation programs in primary care settings that linked patients to primary care services, specialist care, and community-based health and social services (CBHSS), summarised in [NIHR evidence review](https://evidence.nihr.ac.uk/alert/care-navigation-is-being-widely-adopted-in-primary-care-but-in-varying-ways/) |
| Provider outcomes:   - feelings of satisfaction, increased communication among primary care providers and community services and among providers. - Increased knowledge and skills; increased trust between navigators and physicians as well as patients and their attorneys.   improved care coordination and follow up. | | |  |
| Outcomes for navigators: job promotion, professional redevelopment | | |  |
| Health system outcomes**:** reduction in emergency room and/or hospital use and prevention of premature institutionalization for older adults. | | |  |

### 3 Evaluability Assessment process

We used evaluability assessment methods to develop the evaluation design (Leviton, Khan et al. 2010, Craig and Campbell 2015). Evaluability assessment (EA) is a rapid, systematic, and collaborative way of deciding whether and how a programme or policy can be evaluated, and at what potential cost. We conducted two workshops with SN stakeholders to ascertain their understanding of how the SN was intended to work, how it might lead to health outcomes and how it may be evaluated.

Workshop participants included staff from South Tyneside Council, local third sector organisations, and South Tyneside Homes including Social Navigators. We allowed the workshop format to evolve to take account of feedback from the preceding workshop, and to enable stakeholders to shape the approach to evaluation.

Table 2 Summary of workshop dates & agenda

| **Workshop** | **Date** | **Agenda** |
| --- | --- | --- |
| 1 | 19 January 2023 | Theory of Change development; understanding the Social Navigators project |
| 2 | 6 March 2023 | Prioritisation of health and wellbeing related outcomes, development of evaluation questions, data considerations and evaluation options |

#### Workshop 1

The first SN EA workshop aimed to provide the evaluation team with an overview of the SN programme and aim, how SN was expected to bring about change, and what success would look like. A Theory of Change approach was used to clarify the intervention aims and desired outcomes. This was important to establish as social navigators carried out many different activities to support a wide range of needs. While initially focused on finance-related outcomes, the project noted that a wide variety of outcomes not previously anticipated were also being addressed. Stakeholders established that they were keen for the evaluation to show that SN had impacted on health and wellbeing outcomes for servicer users even though SN was not explicitly designed for changes in users in terms of health and wellbeing, Stakeholders also expressed an interest in an economic evaluation of the project.

#### Workshop 2

The second EA workshop focused on getting the Theory of Change signed off (Figure 1). Since SN was not initially intended to impact on service users health and wellbeing, a range of health and related outcomes extracted from the relevant literature was ranked and the results discussed. These were inserted into the signed-off theory of change so that they theory of change could be used to understand unintended project health and wellbeing outcomes (Figure 2). Key evaluation questions were then discussed, as well as data issues (like sources and collection methods) in relation to the evaluation design. Health economic evaluation considerations were also discussed by workshops participants as well as possible evaluation options.

#### Theory of Change

The SN Theory of Change (ToC) was co-developed with workshops participants and signed off in workshop 2. See Figure 1 below.

Figure 1 Social Navigators Theory of Change

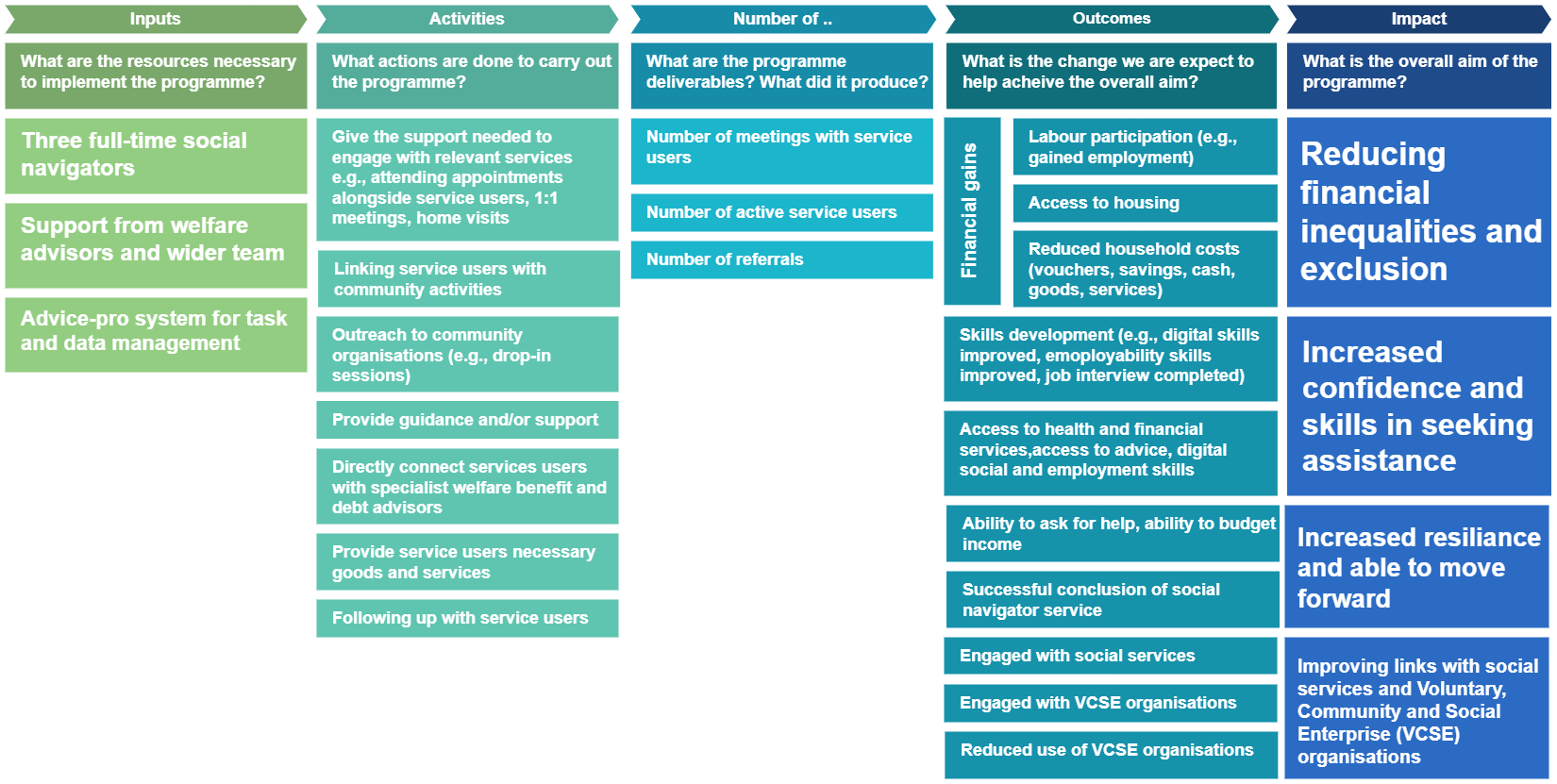

A second theory of change was also developed in workshop 2 with unintended health and wellbeing outcomes included. See Figure 2 below.

Figure 2 Social Navigators Theory of Change with Health & Wellbeing Outcomes

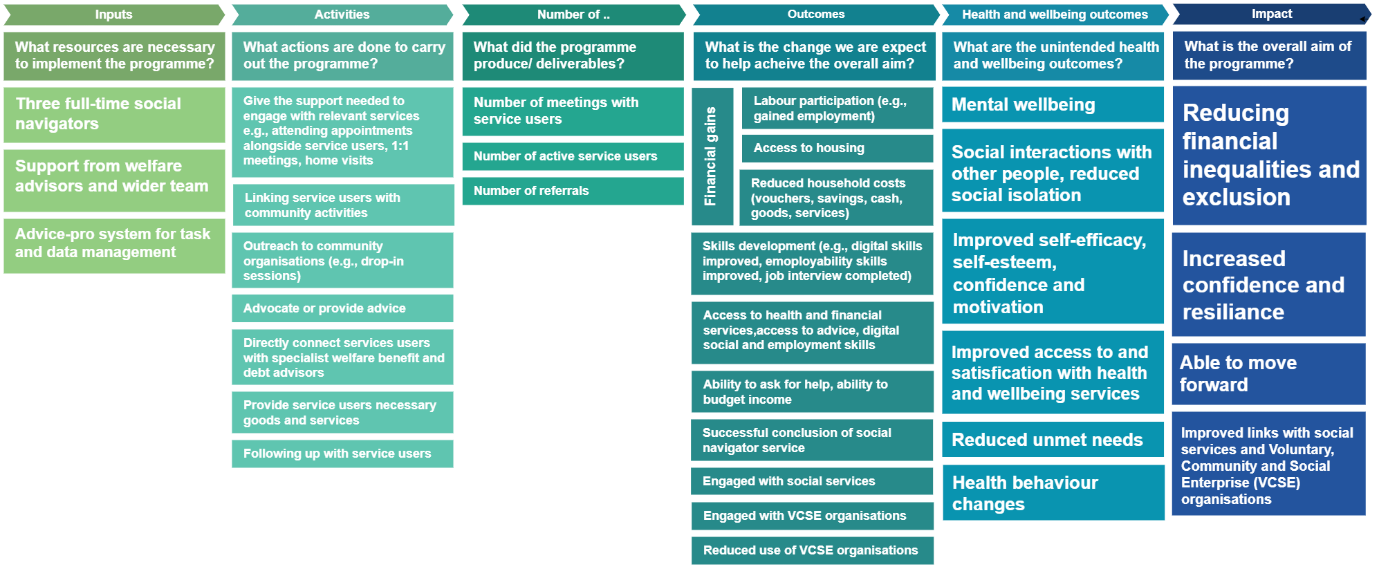

### 4 Evaluation design considerations

#### Aim & evaluation questions

The overall aim of the SN evaluation is to understand how and why improvement in financial gains for project participants lead to better health and wellbeing.

The primary evaluation questions are:

1) What is the relationship between financial gains and health and wellbeing changes for service users? (what is the ToC?)

2) How does the SN lead to better
   a. Improved confidence & motivation
   b. Improved mental wellbeing
   c. Decreased loneliness & social isolation
   d. increased opportunities

The secondary evaluation question is:

1) What is the impact of SN on other services to better support their own clients?

#### Data

Existing data collected by SN includes:

1. Existing service monitoring data collected by the Social Navigators since the start of the project on the interactions with clients at 12 week intervals (after the initial assessment at 6 weeks). This includes data on clients’ engagement with other services, such as welfare support, adult social care, CAB and the job centre, and data on financial gains for clients (child benefits, council tax rebate, debt relief order, food & utility vouchers).
2. Anecdotal evidence in the system on increased confidence (completed job interviews), skills acquisition (digital, social and employment) and wellbeing perceptions (keeping families together, mental health?).
3. Baseline, mid-point and end point data since September 2021 on eight outcome indicators (ability to ask for help, access to advice, health and financial services, ability to budget income, digital social and employment skills) using scale questions (1-10?) for 143 people who have not engaged previously in benefits advice.

#### Evaluation approach

In a typical evaluation of the impact of a service or policy, an effectiveness study is adopted to understand to what extent does the intervention produce the intended outcome(s) in

real-world settings. Effectiveness studies focuses on identifying whether or not interventions ‘work’ based primarily on average estimates of effect. This is not the case of SN. Here, stakeholders have told us in the workshops that they think financial gains by servicer users may have also led to health and wellbeing outcomes. While there is anecdotal evidence of this, especially from social navigators themselves, there is little understanding about why and how this happens. This means that an alternative evaluation approach is required.

##### Theory-based perspective

As recommended by the updated MRC framework for developing and evaluating complex

Interventions (Skivington *et al* 2021), a theory-based evaluation seeks to provide evidence on the processes through which interventions lead to change in outcomes and what prerequisites may be required for this change to take place, thus exploring how and why they bring about change. This takes into account of context, and often explore more than one single theoretical account of how the intervention may work. Such approaches to evaluation aim to broaden the scope of the evaluation to understand how an intervention works and how this may vary across different contexts or for different individuals. Research from this perspective can generate an understanding of how mechanisms and context interact, providing evidence that can be applied in other contexts. Process evaluation designs are most appropriate for theory-based approaches (Moore *et al* 2015).

##### Economic consideration

Stakeholders in the EA workshops also said that they were interested in an economic evaluation of SN to ascertain whether the benefits, including health and wellbeing outcomes, justify the costs of delivering the intervention. There are two traditional approaches to this kind of evaluation. Cost Benefit Analysis (CBA) measures if the benefits of an intervention in monetary terms exceed the costs of the intervention. Cost Consequence Analysis (CCA) allows the costs and outcomes of the intervention to be presented in a descriptive format leaving the decision maker to form a value judgement on whether benefits justify the costs of delivering the intervention.

An alternative way is the ‘social value’ approach. Social value is “the quantification of the relative importance that people place on the changes they experience in their lives accounting for the broader human and societal factors that result from an intervention.” (Ashton, Schroder-Back et al. 2020). This is a departure from traditional ‘value for money’ approaches which recognises the potential added value of public health interventions on an individual’s health and well-being. Hence it is also important to capture the wider social value of interventions, services and policies.

### 5 Evaluation options

Four options are presented in this chapter for starting with the cheapest and most basic, and building on additional evaluation activity:

- Option 0 involves no change or additional evaluation approach and relies on relevant and existing Monitoring & Evaluation (M&E).
- Option 1 supplements Option 0 with a mixed-method summative process evaluation.
- Option 2 is similar to Option 1 but is a formative evaluation approach.
- Option 3 is similar to Option 2 but includes a health economic evaluation to address the secondary evaluation questions.

See Table 3 for an overview

Table 3 Overview of SN evaluation options

| **Evaluation Questions** | **Evaluation Design** | **Elaboration & Data Collection Tools** | **Pros** | **Cons** |
| --- | --- | --- | --- | --- |
| **What is the impact of the project on client’s health and wellbeing?** | **Option 0 (reference option)**  Existing Monitoring & Evaluation (M&E) | - Existing project monitoring data collection (‘8 indicators) | - No additional cost or resources required; can meet funding deadline | - Unable to provide more detailed and contextual data on individual clients |
|  | **Option 1**  Mixed-Method Summative Process Evaluation | - Mixed method summative process evaluation of intervention (eg., semi-structured interviews & existing project monitoring data). This can be split into two phases provided an embedded researcher is available to assist with access to monitoring data | - Can provide more detailed and contextual data on individual clients | - Takes more time and resources to carry out. Unable to meet funding deadline |
|  | **Option 2**  Mixed-Method Formative Process Evaluation | - As above with mid-point check in for dissemination of preliminary findings to support project learning from secondary monitoring data analysis | - Can provide more detailed and contextual data on individual clients - Can support project learning and meet funding deadline (Jul/Aug) | - Inference from mid-point reporting will be drawn on monitoring data only |
|  | **Option 3**  Mixed-Method Formative Process & Health Economic Evaluation | - As above with additional Health Economic Evaluation (Cost Benefit Analysis?) | - As above and can provide further health economic evaluation findings | - Most resource-intensive - Requires appropriate and accessible data |

The rest of this chapter describes each option in detail.

#### Option 0: Existing M&E

This is the reference option. Option 0 involves no change or additional evaluation approach and relies on relevant and existing Monitoring & Evaluation (M&E) system. This includes:

4. Existing service monitoring data collected by the Social Navigators since the start of the project on the interactions with clients at 12 week intervals (after the initial assessment at 6 weeks). This includes data on clients’ engagement with other services, such as welfare support, adult social care, CAB and the job centre, and data on financial gains for clients (child benefits, council tax rebate, debt relief order, food & utility vouchers).
5. Anecdotal evidence in the system on increased confidence (completed job interviews), skills acquisition (digital, social and employment) and wellbeing perceptions (keeping families together, mental health?).
6. Baseline, mid-point and end point data since September 2021 on eight outcome indicators (ability to ask for help, access to advice, health and financial services, ability to budget income, digital social and employment skills) using scale questions (1-10?) for 143 people who have not engaged previously in benefits advice.

We include Option 0 as a reference option. As discussed in the workshops, existing M&E systems do not address SN evaluation requirements.

#### Option 1: Mixed-method Summative Process Evaluation

Option 1 supplements Option 0 with a mixed-method summative process evaluation. This can provide more detailed and contextual data on individual clients but will take more time and resources to carry out. It will also be unable to provide findings to support end-of-funding reporting deadline.

This option involves conducting semi-structured interviews with clients to explore in more detail the perceived health and wellbeing benefits of the Social Navigator project. Clients will be sampled from the existing monitoring data to represent different referral pathways, varying lengths of support received from Social Navigators and a range of social-economic characteristics (age, gender, marital status, income). Some of the qualitative data might be collected through small focus groups. Findings will be reported at the conclusion of the evaluation.

Contribution analysis will be used to process the data. Data collected will be used to iteratively refine the service’s theory of change at the end of the evaluation.

#### Option 2: Formative Process Evaluation

Option 2 is similar to Option 1 but is a formative evaluation approach with one check-in time point around the mid-point of the evaluation to provide timely data for programme learning & adaptation as well as meeting end-of-funding reporting deadline. This formative option allows for several iteration of SN’s theory of change.

#### Option 3: Mixed-Method Formative Process & Health Economic Evaluation

Option 3 is like Option 2 but includes a health economic evaluation to address the secondary evaluation questions. The social value of wellbeing for clients engaged in the social navigators’ project by applying the HACT Wellbeing values (<http://hact.org.uk/publications>) (Cheetham, Van der Graaf et al. 2018). We will perform regression analysis to estimate the relationships between subjective wellbeing and the various outcome variables included in the value bank.

### 6 Recommendation

Option 3 is the recommendation evaluation design because it can provide more detailed and contextual data on individual clients as well as using existing data collected by the programme. It can also provide some health economic evaluation. To suppot project learning as well as to meet funders’ deadline requirements, we propose the following evaluation design organised around 3 work packages (WP) based on Option 3:

#### WP1: Secondary data analysis of existing monitoring data

We will analyse 3 sources of existing service monitoring data collected by the Social Navigators since the start of the project:

- Interactions with clients at 12 week intervals (after the initial assessment at 6 weeks). This includes data on clients’ engagement with other services, such as welfare support and data on financial gains for clients.
- Anecdotal evidence in the system on increased confidence (completed job interviews), skills acquisition (digital, social and employment) and wellbeing perceptions (keeping families together, mental health.
- Baseline, mid-point and end point data since September 2021 on eight outcome indicators (ability to ask for help, access to advice, health and financial services, ability to budget income, digital social and employment skills).

Analysing these three types of existing data (interactions, staff assessments, and outcomes), will help to identify the impact of the project on the financial stability of clients and explore available indicators for health and wellbeing impacts. The data will also help to map referral pathways for clients that we can follow-up with referred-to organisations to explore available data within their organisations about the support provided and benefits experienced by referred clients.

Potential gaps and missing data will inform primary data collection in WP2 and suggestions for inclusion of additional indicators related to health and wellbeing in the monitoring system. The rapid literature review has already identified several potential indicators that could be used to measure relevant health and wellbeing outcomes more systematically with clients. These potential indicators could be piloted with a sample of clients in WP2.

#### WP2: Semi-structured follow up interviews with clients to explore and develop health and wellbeing outcome indicators

We will conduct semi-structured interviews and small focus groups to explore in more detail the perceived health and wellbeing benefits of the Social Navigator project. Clients will be sampled from the existing monitoring data to represent different referral pathways, varying lengths of support received from Social Navigators and a range of social-economic characteristics (age, gender, marital status, income).

Potential indicators for health and wellbeing outcomes will be piloted with participants as part of the interviews to test their feasibility in terms of clients’ understanding and ability to complete the questions, and how well they feel these questions represent the health and wellbeing outcomes they experienced from the social navigator projects.

Contribution analysis will be used to process the data. Data collected from WPs 1 & 2 will be used to iteratively refine the service’s theory of change (co-developed with stakeholders during evaluability assessment workshops).

#### WP3: Health economic modelling of health and wellbeing outcomes

Based on insights from WP1 and 2, we will calculate the social value of wellbeing	for clients engaged in the social navigators’ project by applying the HACT Wellbeing values (<http://hact.org.uk/publications>), suggested by Cheetham et al. (2018).
