## Supplementary File 2 for "Improving local authority financial support services for users with complex health needs: a mixed-method economic evaluation of Social Navigators in South Tyneside, UK"

Supplementary File 1.

Evaluation framework for social prescribing applied to our study

| **Box 1 Recommendations for social prescribing evaluation (Elliott et al. 2022)** | **PHIRST Fusion Social Navigators service evaluation (Van der Graaf et al. 2024)** |
| --- | --- |
| 1. Apply a mixed-methods design to produce an evaluation which captures the impact of social prescribing at multiple levels. | Mixed methods design combining analysis of service monitoring data with semi-structured interviews with service user, conducted by peer research, and a social return on investment analysis to capture impact at user, service and local authority level. |
| 2. Where possible, design social prescribing evaluations iteratively, so that each stage can build upon the previous stage so knowledge can be accumulated and the evidence base can continue to grow. | Evaluation designed iteratively with results from service monitoring data informing interview questions and matching of outcome data from the UK Social Value bank. |
| 3. Undertake a mapping exercise to identify all stakeholders for a social prescribing evaluation. Involve stakeholders from the outset to coproduce the study design and materials. | Stakeholders were involved from the outset in coproducing the study through evaluability assessment workshops with relevant stakeholders identified through a mapping exercise with the local authority at the start of the process. |
| 4. Involve stakeholders in the interpretation, analysis and dissemination of findings so that the evaluation is grounded in the real world and findings can be translated back into practice, to make a difference to people involved in social prescribing. | Social Navigators were involved as peer researchers to support data collection, interpretation and dissemination. An embedded research further support access to data, interpretation and dissemination with direct links to practice and decision-makers that achieved recommissioning of the service and ongoing work to map and align support services across the Borough. |
| 5. Involve members of the public throughout social prescribing evaluation in a meaningful way. Follow the UK Standards for Public Involvement and report public involvement when disseminating findings. | Four members of the public were involved in the evaluability assessment workshops to design the study. Two members of the public are members of the independent advisory group for PHIRST Fusion to provide strategic oversight on project progress and dissemination. Their involvement is reporting in this paper and the research report (add link). |
| 6. Take time at the start of the evaluation, before the study design is determined, to understand the social prescribing intervention or service that is going to be evaluated. Identify the aims, objectives, participants, context, setting, activities, processes that are involved. | To understand the Social Navigators service form the start, our PHIRST Fusion Knowledge Exchange Broker scoped our evaluation needs and aims with local authority stakeholders in online meetings. This informed the design and delivery of two Evaluability Assessment Workshops to gauge their wider stakeholder understanding of how the SN service was intended to work, how it might lead to health outcomes and how it may be evaluated, resulting in a Theory of Change. Workshop participants included staff from South Tyneside Council, South Tyneside Homes, local third sector organisations, and public members who have engaged with the Social Navigators. |
| 7. Align the evaluation with the social prescribing intervention so that the evaluation can answer questions that are relevant to the intervention and to stakeholders. | The EAW ensured that our evaluation aligned with the intervention and questions that were relevant to local stakeholders. |
| 8. Seek advice or use an evaluation framework to inform evaluation decision making. This will maximise data quality, and ensure a consistent approach which can be compared with other similar evaluations. | Our evaluation approach was informed by the suggestions and preferences from local stakeholders, including utilising available service monitoring data, collecting insights from service users using peer researchers, and enhancing this data by conducting a Social Return On Investment analysis that could be repeated to inform future monitoring and evaluation of the service. We compared our evaluation approach to the 15 recommendations developed by Elloitt et al. (2022) to check consistency and share learning about the application of these recommendations in a particular context. |
| 9. For rigorous evaluations of social prescribing, remove the burden on link workers and use independent researchers to collect data at the appropriate time point. | Social Navigators were keen to be involved in the data collection and interpretation; their involvement greatly enhanced the access to and quality of data. We supported them in their role as peer researchers by providing training, equipment, recruitment and information materials and organising debrief meetings after interviews. The overall evaluation was conducted by independent researchers from PHIRST Fusion, removing the burden from the local authority. However, we have extended recommendations 3 and 4 to include data collection with stakeholders to create more opportunities for co-production and translation into practice. |
| 10. Provide sufficient funding for social prescribing evaluations, to ensure that they can be undertaken rigorously, without bias, to address gaps identified by services or in the literature. | Funding was provided by NIHR PHIRST, which included a full-time research associate, a 0.4FTE embedded researcher, and a 0.2FTE health economist, to ensure a rigorous evaluation with sufficient resources. |
| 11. Integrate mixed-methods findings to generate a more in-depth, nuanced understanding of social prescribing, how it works, for whom and in what context. | We integrated our mixed methods findings to provide an in-depth understanding of the vulnerable population served by the Social Navigators, how they supported users to access services and financial gains and how this support impacted on their health and wellbeing in South Tyneside. |
| 12. Triangulate findings from multiple data sources and different perspectives to generate a more in-depth, nuanced understanding of social prescribing, how it works, for whom and in what context. | We triangulated our findings from the secondary data analysis, interviews and SROI analysis, combining perspectives from service users, social navigators and national survey data to generate a more nuanced understanding of social prescribing with a focus on non-clinical setting and the wider determinants of health/ financial health. |
| 13. When using mixed-methods or conducting a multicomponent study, produce an overarching commentary or narrative, explaining the links between the different components and identifying remaining gaps for future research. | We produced on overarching narrative of our mixed methods to explain the links between out different components and identified clear recommendation for future development and research. |
| 14. Provide in-depth descriptions of methods used and decisions made to facilitate judgements about the rigour and quality of the study, and to enable the study to be replicated in different contexts. | We have described our use of methods in-depth in our Methods section, including our various mechanisms for co-production, and the decisions we made in our data analysis to demonstrate rigour and quality and enable replication of the study in other contexts. |
| 15. Report good practice, strengths, successes, failed approaches and methods to mitigate challenges in social prescribing evaluation to support future evaluators. | We have reported on our strength and limitations, include missing data and gaps in the service monitoring data, to strengthen future data collection and evaluation. For example, we made recommendations for better aligned between scored outcomes for service monitoring and available outcomes in the HACT social value database. |
