## Supplementary File 3 for "Improving local authority financial support services for users with complex health needs: a mixed-method economic evaluation of Social Navigators in South Tyneside, UK"

Table F3.1. Ongoing gains sources, and median gain per source

| **Gain description** | **Frequency** | **Mean ± SD** | **Median (IQR)** |
| --- | --- | --- | --- |
| Attendance Allowance High | 4 | £4854 ± £300 | £4732 (£4659 to £4926) |
| Child Benefit | 1 | £1267 ± £0 | £1267 (£1267 to £1267) |
| Council Tax Support | 12 | £740 ± £292 | £808 (£693 to £901) |
| Discretionary Housing Payment | 6 | £696 ± £367 | £572 (£418 to £772) |
| Energy Savings | 1 | £150 ± £0 | £150 (£150 to £150) |
| Housing Benefit | 2 | £5270 ± £916 | £5270 (£4947 to £5594) |
| LCWRA | 4 | £4359 ± £215 | £4251 (£4251 to £4359) |
| Northumbrian Water Scheme - Weekly/Monthly | 1 | £576 ± £0 | £576 (£576 to £576) |
| Occupational Pension | 1 | £6133 ± £0 | £6133 (£6133 to £6133) |
| Other Savings Measure | 3 | £212 ± £157 | £150 (£123 to £270) |
| Pension Credit | 4 | £6891 ± £2951 | £7132 (£5480 to £8423) |
| PIP Daily Living Enhanced Rate | 14 | £4833 ± £150 | £4805 (£4805 to £4805) |
| PIP Daily Living Standard | 15 | £3291 ± £143 | £3216 (£3216 to £3216) |
| PIP Mobility Enhanced Rate | 14 | £3340 ± £38 | £3354 (£3354 to £3354) |
| PIP Mobility Standard Rate | 13 | £1300 ± £56 | £1271 (£1271 to £1271) |
| State Pension | 2 | £8531 ± £3862 | £8531 (£7165 to £9896) |
| Tax Credit | 2 | £6081 ± £2938 | £6081 (£5043 to £7120) |
| UC - Carer's Element | 3 | £1995 ± £43 | £1995 (£1980 to £2010) |
| UC Child Disability Element | 1 | £1595 ± £0 | £1595 (£1595 to £1595) |
| Universal Credit | 12 | £6771 ± £5716 | £4047 (£3180 to £10244) |

Table F3.2. One-off gain sources, and median gain per source

| **Gain description** | **Frequency** | **Mean ± SD** | **Median (IQR)** |
| --- | --- | --- | --- |
| Attendance allowance (backdated) | 3 | £885 ± £435 | £722 (£638 to £1050) |
| Tax Credit | 2 | £3441 ± £2204 | £3441 (£2662 to £4221) |
| Children in Need | 1 | £214 ± £0 | £214 (£214 to £214) |
| Community Shop | 1 | £1040 ± £0 | £1040 (£1040 to £1040) |
| Council Tax Energy Rebate | 8 | £140 ± £19 | £150 (£142 to £150) |
| Council Tax Refund | 1 | £112 ± £0 | £112 (£112 to £112) |
| Debt Managed | 12 | £2832 ± £3581 | £1121 (£753 to £2957) |
| Debt Relief Order Fee | 4 | £90 ± £0 | £90 (£90 to £90) |
| Debt Written Off | 15 | £5701 ± £6092 | £3689 (£1138 to £7717) |
| Discretionary Housing Payment - Lump Sum | 2 | £188 ± £6 | £188 (£186 to £190) |
| Energy Saving Lightbulbs | 22 | £5 ± £0 | £5 (£5 to £5) |
| Energy Savings | 21 | £13 ± £9 | £10 (£10 to £10) |
| Energy Voucher | 34 | £57 ± £29 | £49 (£49 to £49) |
| Financial gain - Charitable payment | 8 | £210 ± £170 | £145 (£88 to £288) |
| Financial gain other | 24 | £2113 ± £3745 | £316 (£99 to £2328) |
| Groundworks | 1 | £25 ± £0 | £25 (£25 to £25) |
| Household Support Fund | 42 | £126 ± £30 | £100 (£100 to £150) |
| Housing benefit backdate | 1 | £2208 ± £0 | £2208 (£2208 to £2208) |
| Key 2 Life Xmas Presents To Client | 5 | £30 ± £0 | £30 (£30 to £30) |
| LEAP White Goods | 8 | £476 ± £158 | £560 (£302 to £602) |
| Lucie's Pantry Shopping | 30 | £87 ± £199 | £13 (£13 to £13) |
| LWP Community Care Grant | 24 | £428 ± £266 | £394 (£245 to £530) |
| LWP Food/Utility Vouchers | 12 | £49 ± £23 | £40 (£35 to £60) |
| Nexus Taxi Card | 1 | £90 ± £0 | £90 (£90 to £90) |
| Other Energy Saving Products | 4 | £13 ± £5 | £15 (£13 to £15) |
| Other Grant Award | 18 | £158 ± £141 | £90 (£50 to £238) |
| Pension Credit Backdate | 3 | £2649 ± £2675 | £2242 (£1221 to £3873) |
| PIP backdated | 24 | £2158 ± £962 | £2113 (£1459 to £2793) |
| Rent Refund | 4 | £606 ± £168 | £609 (£492 to £724) |
| The Green Doctor Energy Grant | 2 | £49 ± £0 | £49 (£49 to £49) |
| UC Child Disability Element (backdate) | 1 | £516 ± £0 | £516 (£516 to £516) |
| Universal Credit Backdate | 7 | £762 ± £443 | £530 (£476 to £985) |
| Warm Home Discount | 5 | £140 ± £0 | £140 (£140 to £140) |
| White Goods | 11 | £312 ± £156 | £253 (£204 to £385) |
| Wise Steps Heat Fuel Team | 21 | £150 ± £0 | £150 (£150 to £150) |
